# Insula Network Microstructural Injury Links Low-Level Blast Exposure to Clinical Depression in Military Special Operations Forces Soldiers

**DOI:** 10.64898/2026.05.12.26353040

**Authors:** Cory McEvoy, Adam Crabtree, Daniel Murray, Mohamed Omer, Josh W. Rodriguez, Trinity Charles, Tami Wolden-Hanson, Donghoon Lee, Todd Richards, Ronald G. Thomas, Elaine Peskind, Jason P. Mihalik, James S. Meabon

**Affiliations:** United States Army Special Operations Command, Fort Bragg, NC, USA; Stanford University School of Medicine, Stanford, CA, USA; CU Anschutz Center for COMBAT Research, Department of Emergency Medicine, University of Colorado School of Medicine, Aurora, CO, USA; School of Medicine, University of Nevada, Reno, NV, USA; VA Northwest Mental Illness Research, Education, and Clinical Center (MIRECC), VA Puget Sound Health Care System, Seattle, WA, USA; Rodent Metabolic and Behavioral Phenotyping Core, R&D, VA Puget Sound Health Care System, Seattle, WA, USA; Department of Radiology, University of Washington, Seattle, WA, USA; Alzheimer’s Disease Cooperative Study, University of California San Diego, La Jolla, CA, USA; Division of Biostatistics, Department of Family Medicine & Public Health, University of California San Diego, La Jolla, CA, USA; Department of Psychiatry and Behavioral Sciences, University of Washington, Seattle, WA, USA; Matthew Gfeller Center, Department of Exercise and Sport Science, University of North Carolina at Chapel Hill, Chapel Hill, NC, USA

**Keywords:** Occupational blast exposure, neural resilience factors, subconcussion, career risk, military, low-intensity blast, resiliency, neurodegeneration

## Abstract

Special Operations Forces (SOF) sustain repeated low-level blast (LLB) exposures; while most remain resilient, a subset develop depression, sleep disruption and reduced wellbeing that threaten readiness. We asked whether “sub-concussive” LLB chronically injures an insula-centered cortico-striato-thalamic network and whether network architecture explains divergent outcomes. In a mouse model parameterized to SOF blast exposure monitoring data, ninety 3-psi blasts over three weeks produced persistent diffusion and connectivity deficits across insular, striatal, pallidal and thalamic nodes, accompanied by tauopathy, neuroinflammation, vascular amyloid, and durable sleep and metabolic abnormalities. In SOF Soldiers, measures of cumulative LLB exposure predicted right insula-striatal diffusion/neurite disruption and increased depression risk. Interventional multi-mediator modeling showed that right insula-striatal microstructural injury mediated the effect of LLB to increase depression risk, while moderator screening identified features that amplify or buffer this mediation, defining risk and resilience zones. These findings enable precision blast-medicine integrating exposure dose, circuit biomarkers and moderator profiles.

## Introduction

Low-level blast (LLB) overpressure has become a ubiquitous feature of modern military training and operations, particularly in communities that rely heavily on explosive breaching and weapons systems. Emerging monitoring programs, including the Department of Defense/War’s CONQUER initiative and related blast-surveillance efforts, demonstrate that Service members can accumulate hundreds to thousands of “sub-threshold” overpressure events over a career^1,2^. These exposures have complex, weapon-specific profiles of peak pressure and impulse that are rarely captured by concussion-focused surveillance. As a result, concern has shifted from isolated high-level blast injuries toward the cumulative effects of repetitive LLB exposures that fall below current 4 psi occupational limits but may still perturb brain structure, physiology, and behavior over time^3,4^.

Across human cohorts, LLB exposure has been repeatedly linked to neuropsychiatric symptoms that are directly relevant to warfighter readiness, including depression, irritability, cognitive slowing, and sleep disturbance. Early descriptive work in breachers and range staff found that chronically blast-exposed personnel report more headaches, post-concussive symptoms, and lower energy than matched controls^5,6^. Subsequent observational studies in combat Veterans and training populations show that blast severity and cumulative blast exposure predict higher depressive and neurobehavioral symptom burden even after accounting for PTSD diagnosis and mild TBI history, suggesting that blast itself can exert an independent influence on mood and function^7^. Although most individuals with blast exposure never develop severe psychiatric sequelae, the possibility that repeated LLB could incrementally increase depression burden has heightened urgency for mechanistic models that connect realistic blast patterns to specific brain injuries and clinically meaningful outcomes^8,9^.

Recent work in Special Operations personnel has begun to reveal network-level and neuroimmune correlates of repeated blast exposure but has not resolved how these changes relate to clinically significant depression. In experienced breachers or other high-risk military occupational specialties, studies have described altered cortical thickness^10,11^, decreased white-matter integrity^12^, reduced default-mode network activation^11^, neuroinflammatory signatures in brain-derived extracellular vesicles^13,14^, and dysregulated immune-related gene expression^15^.

Higher generalized blast exposure values derived from detailed blast histories have been associated with cortical thickness changes in the rostral anterior cingulate cortex, altered functional connectivity in salience and executive networks, tau deposition and neuroinflammation, and worse health-related quality of life and neurobehavioral symptom burden^11,16,17^. Parallel work in blast-mTBI Veterans has identified basal ganglia metabolic alterations that mediate executive dysfunction, underscoring vulnerability of cortico-striato-thalamo-cortical circuits to blast^18,19^.

A major translational limitation is that many rodent shock-tube paradigms rely on single or few exposures at moderate or high overpressures (>10-20 psi), generating clear neuropathology and anxiety- or depression-like behaviors but operating in a pressure and dosing regimen more typical of high-level blast or frank TBI than routine training^20,21,22^. These models consistently demonstrate chronic tau phosphorylation, neuroinflammatory activation, white-matter injury, and persistent affective and cognitive deficits after repeated blasts, but the mismatch in magnitude, frequency, and cumulative dose relative to operational LLB has raised questions about relevance for warfighter brain health^4,3^. Recent concept papers and scoping reviews argue that preclinical studies must be parameterized from monitored exposures in weapons training to approximate the heterogeneous yet predominantly sub-threshold overpressure environment encountered in military training^1,2,3,23,24^.

Mechanistically, sub-threshold does not imply biologically inert. Blast waves combine peak pressure with rise time, positive-phase duration, and impulse^25^; for lower-peak exposures, impulse and repetition plausibly focus loading at vascular-parenchymal interfaces, among other places^26,27,28^. The neurovascular system introduces pronounced impedance contrasts among blood, vessel wall, cerebrospinal fluid, and parenchyma, raising the possibility that repetitive LLB perturbs vulnerable neurovascular-adjacent white matter and hub circuits that integrate interoceptive state with action selection. The insula is a candidate hub in this framework, connecting central autonomic network (CAN) nodes with basal ganglia and thalamus to regulate arousal, salience, sleep, and affect.

In the present study, we developed an empirically parameterized cross-species framework centered on an insula-basal ganglia circuit. First, we built a mouse model of repetitive 3 psi LLB (below the 4 psi threshold) designed to approximate a high-tempo weapons training burden^1,29^ and tested whether it produced chronic injury in an insula-centered cortico-striato-thalamic network together with sleep and metabolic alterations linked to depression. Second, in U.S. Army SOF Soldiers, we combined diffusion and microstructural imaging of insula-striatal and CAN white matter with validated measures of depression, wellbeing, and sleep, using time in Low-Level Blast Occupation as a practical surrogate for cumulative LLB burden within our established characterization of this cohort^15,29^. Finally, we applied interventional multi-mediator (IMM) and moderation analyses to test whether diffuse right insula-basal ganglia injury mediates the association between cumulative LLB and PHQ-9-defined clinical depression risk, and whether network features help explain why individuals with similar exposure histories exhibit heterogeneous outcomes.

## Methods

### Mouse Model

#### Animals

A total of 76 three-month-old male C57BL/6J mice (Strain 000664; Jackson Laboratory, Sacramento, CA) were used in this study. All procedures were approved by the Veterans Affairs Puget Sound Health Care System Institutional Animal Care and Use Committee (protocol no. ACORP0963) and conducted in accordance with institutional and national guidelines for the care and use of laboratory animals and with ARRIVE 2.0 reporting recommendations. Mice were confirmed to be specific pathogen-free on arrival. At the start of the experimental procedures, mice weighed 28-30 grams and were free of overt signs of illness.

Mice were housed in an AAALAC-accredited specific pathogen-free facility at the Veterans Affairs Puget Sound Health Care System (Seattle, WA, USA) in groups of 3-5 per cage, with cob bedding and standard environmental enrichment. The housing room was maintained at 20-26 °C and 30-70% relative humidity on a 12-h light-dark cycle (lights on 6 am to 6 pm, PST) with ad libitum access to standard rodent chow (PicoLab 5053 irradiated diet, LabDiet, Gray Summit, MO) and water. Animals were monitored at least twice daily for general health and signs of distress throughout the study.

Mice were cage-randomized to low-level blast (LLB) or sham exposure groups (44 and 32 animals per group, respectively). Investigators performing MRI, behavioral testing, neuropathology and related analyses were blinded to group allocation. No animals were excluded from the analysis.

#### Mouse model of Career Low-Level Blast Exposure

Mice assigned to the LLB group underwent six sequential LLB exposures delivered once per day, Monday-Friday while isoflurane anesthetized (2.5% in 1 L min⁻¹ O₂) for a single ∼14-min session as established^24^. During LLB exposure, mice were positioned with their dorsal surface against a rigid metal-mesh gurney and secured at the wrists, ankles and torso, allowing the entire ventral surface to face the oncoming overpressure wave. The mean (± s.e.m.) LLB overpressure characteristics of 1071 randomly selected events were 3.01 ± 0.007 pounds per square inch (psi; ∼269 cumulative psi with peak single event pressures ranging from 3.38-4.45 psi), 0.935 ± 0.036 ms and 1.82 ± 0.00002 psi·ms (∼164 cumulative psi·ms) Sham-exposed mice underwent identical anesthesia and handling procedures without exposure to the overpressure wave. All animals survived to the planned 6-month post-LLB/sham study endpoint, at which time they were euthanized by lethal overdose of sodium pentobarbital (210 mg/kg, intraperitoneal), followed by transcardial perfusion with 1 M phosphate buffered saline (PBS) and exsanguination in accordance with AVMA Guidelines for the Euthanasia of Animals.

#### Mouse Behavioral Analyses

<u>Sleep</u>. Sleep was assessed non-invasively using an automated piezoelectric sleep-wake scoring system for rodents (PiezoSleep; Signal Solutions, Lexington, KY, USA). Two months after the final TBI or sham procedure, mice were individually housed in open-floored recording chambers equipped with piezoelectric sensors that detect and classify the previously characterized sleep activity of mice^30^. Animals were allowed to acclimate to the chambers for 2 days before their undisturbed activity was recorded continuously for the subsequent 3 days, following established protocols^31,32^. Sleep bout duration and the sleep-wake ratio were extracted from the raw signals and analyzed using the Sleep Statistics Toolbox (Signal Solutions, Lexington, KY, USA). <u>Metabolic Phenotyping.</u> Metabolic phenotyping was performed using a computer-controlled indirect calorimetry system (CLAMS-HC; Columbus Instruments, Columbus, OH, USA) in the Rodent Metabolic Phenotyping Core at the VA Puget Sound Health Care System. Mice were singly housed for 5 days in home-cage-like chambers (Tecniplast 1250; floor area 580 cm²) within a temperature- and light-controlled cabinet (30 ± 1 °C; 12 h light-dark cycle) with irradiated corncob bedding and environmental enrichment. Food and water were provided ad libitum and mounted on load cells for continuous intake monitoring, and locomotor activity, cage temperature and humidity were recorded throughout. O₂ consumption (VO₂) and CO₂ production (VCO₂) were measured for 20 s, nine times per mouse per hour, using an integrated zirconia O₂ sensor and nondispersive infrared CO₂ sensor, calibrated at the start of each experiment with reference gas mixtures. Incurrent air reference values were sampled after every 16 cages, and the respiratory quotient (RQ) was calculated as VCO₂/VO₂. Data acquisition and analysis were performed using Oxymax software (v5.68; Columbus Instruments, Columbus, OH, USA). <u>Murine Body Composition Measures</u>. Body composition was assessed in unanesthetized mice by quantitative magnetic resonance using an EchoMRI 4-in-1-700 instrument (Echo Medical Systems, Houston, TX, USA) at the VA Puget Sound Health Care System Rodent Metabolic Phenotyping Core.

#### Mouse Diffusion Magnetic Resonance Imaging and Analyses

##### Acquisition

Following euthanasia, mice used for *ex vivo* MRI were were opened at the thoracic cavity and intracardially perfused (1 mL/min), first with 30 mL of 1 M PBS containing 3 mL heparin and 5mM gadolinium followed by 30 mL of 10% neutral buffered formalin containing 5 mM gadolinium (Prohance gadoteridol, Bracco Diagnostics, Princeton, NJ, USA). Mice were then decapitated, defleshed, and the lower jaw was removed. Brains were stored in-skull in 1 M PBS / 5 mM gadolinium / 0.02% sodium azide (4° C), fixed to a depressor, and placed in a 15 mL tube filled with Fomblin Y (Cat. 317926; Sigma-Aldrich, St. Louis, MO, USA). 14T 3D-DTI was conducted using echo-planar imaging (EPI) with repetition time of 500 ms, echo time of 14.4 ms, number of segmentation of 4, field of view of 12.8 x 12.8 x 17 mm, matrix of 128 x 128 x 34, number of averages of 2, diffusion gradient duration of 2.5 ms, diffusion gradient separation of 7.5 ms, diffusion gradient b values of 1000 and 2000 s/mm 2, and number of diffusion directions of 30 and 30.

##### Diffusion MRI preprocessing

Diffusion-weighted MRI data acquired on a 14T small-bore system were preprocessed before network analysis. FSL is a comprehensive library of analysis tools for FMRI, MRI and diffusion brain imaging data^33,34,35^. Raw diffusion images were first corrected for eddy-current-induced distortions and head motion using the eddy tool in the FMRIB Software Library (FSL). Voxelwise diffusion tensor parameters, including fractional anisotropy (FA) and mean diffusivity (MD), were then estimated with FSL’s dtifit utility. These tensor-derived maps formed the basis for subsequent tractography and group-level comparisons between sham- and TBI-exposed mice. Group differences in FA and MD were summarized at the voxel and regional levels using z-score maps derived from sham-TBI contrasts.

##### Registration to the Allen Mouse Brain Common Coordinate Framework

Processed diffusion data were registered to the Allen Mouse Brain Common Coordinate Framework (CCF) version 3.0^36^ to enable anatomically standardized analyses. Registration proceeded in two stages. First, FA maps were linearly aligned to the CCF template using FSL’s FLIRT (affine transformation). Second, non-linear deformations were estimated using Advanced Normalization Tools (ANTs) to refine alignment and minimize residual mismatch. Custom scripts were developed to read the diffusion-derived images and CCF atlas volumes, apply the estimated transforms, and visualize the registered data. The CCF resource comprises an averaged structural template used for registration; an edge map delineating regional boundaries for overlay on structural or diffusion images; and a high-resolution parcellation atlas defining >600 brain regions as a three-dimensional label matrix. This atlas was used both to extract region-of-interest (ROI)-level diffusion measures and to identify the anatomical loci of significant z-scores from MRI- and correlation-based analyses after these maps were transformed into CCF space.

##### Insula-related regions of interest

For the assortativity analysis, we focused on a prespecified set of ROIs putatively involved in insula-centered network connectivity. ROIs were defined directly from the CCF atlas and included bilateral insula, caudate, putamen, globus pallidus, amygdala and entorhinal cortex. The numeric prefixes correspond to the atlas index for each three-dimensional region. Tensor values within each ROI were extracted from the registered diffusion maps for each animal.

##### Probabilistic tractography and network construction

To characterize structural connectivity among the nine insula-related ROIs, we performed probabilistic tractography using FSL’s bedpostx and probtrackx pipelines. Bedpostx was used to model diffusion parameters at each voxel and to estimate the distribution of one or more fiber orientations, thereby accommodating crossing fibers. The resulting parameter estimates were used as input to probtrackx, which generated probabilistic streamlines between all ROI pairs. For each mouse, probtrackx produced an ROI-by-ROI connectivity matrix (the fdt_network_matrix output).

Connection strengths were normalized by the corresponding waytotal values to account for differences in seeding and streamline counts across subjects. The normalized matrices were then used as weighted adjacency matrices for subsequent graph-theoretical analyses.

##### Network metrics and assortativity analysis

Graph-theoretical metrics were computed using the Brain Connectivity Toolbox^37^. For each ROI (node), we calculated assortativity using the degree-based assort1 coefficient, which measures the correlation between the connectivity degrees (or strengths) of nodes at either end of each edge in the network. Positive assortativity indicates a tendency for highly connected nodes to preferentially connect to other highly connected nodes (and weakly connected nodes to connect to similarly weakly connected nodes); whereas, negative assortativity reflects preferential connections between high-degree and low-degree nodes. Assortativity coefficients and node strengths were derived for each subject’s insula-related network and used for group-level comparisons between sham and TBI cohorts.

##### Mouse z-score based Injury mapping

Individual DTI measures were calculated for each CCF brain region by taking the average of the DTI measure within each brain region for each subject and outputting a table in CSV format containing one row for each subject and one column for each brain region. FA and MD z-score maps were calculated using gfortran which compares the sham group (N=7) to the LLB group (N=12). The maps were calculated by looping over each spatial element of the 3D matrix.

#### Neuropathology and Immunohistochemistry

For neuropathological analyses, a random subset of mice used in MRI analyses (N=9 per group) assessed by standard immunohistochemistry. Following MRI, brains were de-skulled and post-fixed in 10% neutral buffered formalin for 24-72h at 4 °C, then embedded in paraffin using standard procedures on a Peloris 2 tissue processor (Leica Biosystems, Deer Park, IL, USA). Paraffin blocks coronally sectioned at the forebrain plane containing basal forebrain, basal ganglia and insular cortex on a microtome at 4-5 μm.

Immunohistochemistry was performed on 5 µm paraffin-embedded mouse brain sections using a semi-automated staining platform (IntelliPATH, Biocare Medical, Pacheco, CA, USA) with heat-induced epitope retrieval (Decloaker, Biocare Medical, Pacheco, CA, USA) in either Diva buffer (pH 6.2; DV2004MM, Biocare Medical; 110 °C for 15 min for IBA1, CD68, and GFAP) or Nuclear Decloacker (pH 9.0; CB911M, Biocare Medical; 95 °C for 15 min for pTau217 or 110 °C for 8 min for AT8), followed by a 10-min room-temperature cool-down, rinse in running water, and equilibration in Tris-buffered saline with Tween-20 (TBST). Slides were processed on IntelliPATH according to manufacturer protocols with antibody-specific reagents, including background blocking (IP FLX Background Punisher, IP974; 5 min, where applicable) and endogenous peroxidase quenching (IP FLX Peroxidizer, IPB5000; 10 min, where applicable), with three TBST rinses between steps; primary antibodies were incubated for 30 min (IBA1, Wako 019-19741, 1:1,000; CD68, DAKO/Agilent M0814, 1:500; GFAP, DAKO/Agilent Z0334, 1:1,000; phospho-Tau pTau217, custom antibody (gift from Dr. Hoffnagle), 1:2,000; AT8, Invitrogen MN1020, 1:800), followed by species-appropriate polymer detection (Mach 3 Rabbit Probe (M3R531) + Mach 3 Rabbit HRP for IBA1; Mouse Secondary Reagent + Universal HRP tertiary (M4U534) for CD68; Mach 3 Mouse Probe (MP530) + Mach 3 Mouse HRP (MH530) for GFAP; and Mach 3 Mouse Probe + Mach 3 Mouse HRP for pTau217 and AT8), with 10-min incubations and TBST rinses after each detection step. Immunoreactivity was visualized using HRP-mediated 3,3ʹ-diaminobenzidine (DAB; IntelliPATH DAB, IPK5010) and counterstained with hematoxylin (1:4; IPCS5006L).

Stained slides were scanned using a Leica Aperio AT2 whole-slide scanner at 10x and exported as high-resolution digital images. Quantitative image analysis was performed using Photoshop 2025 (version 27.1; Adobe, San Jose, CA, USA) using ROI-based intensity-histogram measures. Regions of interest were manually defined for entire basal ganglia, basal forebrain and insula, and thresholds for marker-positive signal were set based on sham control tissue intensity histograms and applied uniformly to all regions in each analysis.

#### Imaging mass cytometry and Analysis

Metal-conjugated antibodies were either purchased directly (Standard Biotools, San Francisco, CA, USA) or generated in-house by conjugation using the MaxPar X8 labelling kit (Standard Biotools) and carrier- and preservative-free antibody formulations, following the manufacturer’s instructions. Formalin-fixed, paraffin-embedded mouse brain sections were processed for imaging mass cytometry according to the manufacturer’s protocol for formalin-fixed paraffin-embedded tissue, with minor modifications. Briefly, sagittal sections containing striatal and basal forebrain regions from a sham-exposed mouse and an LLB-exposed mouse were mounted onto Superfrost Plus slides (Thermo Fisher Scientific, Bothell, WA, USA). DNA was labeled with an iridium intercalator (Ir191/193, 201192B; Standard Biotools) diluted 1:4,000 in Dulbecco’s phosphate-buffered saline (21600-051 in 18.2 MΩ water; Thermo Fisher Scientific) for 30 min at room temperature. Slides were then washed in 18.2 MΩ water, air-dried overnight and stored at room temperature until ablation.

Tissue regions of interest were ablated using a Hyperion imaging system (Standard Biotools) with a 1 μm² laser ablation area. Ablated material was carried by a helium stream into a Helios mass cytometer, where a time-of-flight mass detector quantified the abundance of a user-defined mass range at each rasterized two-dimensional coordinate^38^. The Hyperion instrument was tuned daily according to the manufacturer’s recommendations.

Imaging mass cytometry data were acquired in MCD format and exported as single-channel TIFF images for bulk import into CellProfiler. Single-cell segmentation was performed using a watershed gradient algorithm, with DNA intercalator channels (191Ir, 193Ir) used to define nuclei and a proprietary mix of plasma membrane markers (195Pt, 196Pt, 198Pt; IMC Cell Segmentation Kit, TIS-00001; Standard Biotools) used to delineate cell boundaries. Major neural and non-neural cell populations, including macrophages, astrocytes, neurons, oligodendrocytes, peripheral immune cells and vascular endothelial cells, were validated by manual annotation.

Images were registered to the Allen mouse brain reference atlas^39^ and segmented into anatomical regions using HistoCAT^40^. Neighborhood analyses (proximity radius = 6) were performed in HistoCAT using Barnes-Hut t-SNE and Phenograph with default settings. Images representing single and merged channels were exported as TIFF files using MCD Viewer software (Standard Biotools), imported into Adobe Photoshop 2025 (Adobe Systems, San Jose, CA, USA) and processed with a Gaussian blur filter to reduce speckle noise. For all images that were directly compared within a figure panel, brightness and contrast were uniformly adjusted using a single shared adjustment mask.

### Clinical Participant Measures

#### Military Participants

We conducted a single-site, cross-sectional observational study using data collected during the Assessing & Tracking Tactical (ATTAC) Forces Initiative (2020-2023) at the University of North Carolina at Chapel Hill (UNC Chapel Hill). The study population consisted of 245 male Special Operations Forces (SOF) Soldiers who did not self-report any acute injuries or mTBI related symptoms at the time of assessment (51 new SOF trainees, 113 SOF Soldiers completing their organization’s training pipeline, 70 in-career SOF Soldiers, and 11 retired SOF Soldiers). We operationalize these respective groups as New-, Low- and High-LLB Occupational Exposure groups based on their time in Close Quarters Battle (CQB) training roles. It is relevant to the interpretation of the data to note that despite being “new SOF trainees,” these individuals have typically accrued over 10 years of military service, many of those in Special Forces and have both experience in breaching capabilities and exposure to LLB exposures that surpass most Servicemembers in conventional forces. All procedures were originally approved by the Office of Human Research Ethics at the University of North Carolina at Chapel Hill (UNC) and, subsequently, the US Army Medical Research and Development Command’s Office of Human Research Oversight. Verbal informed consent was obtained from all participants before any study activities were initiated.

#### Clinical Diffusion Magnetic Resonance Imaging and Analyses

##### Diffusion MRI

Neuroimaging was performed at the UNC Biomedical Research Imaging Center using 3T Biograph mMR and 3T MAGNETOM Prisma scanners (Siemens Healthineers, Erlangen, Germany). Imaging protocols were harmonized between scanners by a MR physicist. High-resolution anatomical images were acquired with a three-dimensional T1-weighted magnetization-prepared rapid acquisition gradient-echo (MPRAGE) sequence (inversion time [TI], 900 ms, repetition time [TR], 1900 ms; echo time [TE], 2.26 ms, voxel size 0.5 × 0.5 × 1.0 mm³; field of view 256 mm x 256 mm, slice thickness 1mm, 192 slices). Resting-state functional MRI was obtained while participants lay supine with eyes open, maintaining gaze on a central fixation cross. Blood oxygenation level-dependent (BOLD) signals were measured using T2*-weighted echo planar imaging (TR 2300 ms; TE 27 ms; slice thickness 3.5 mm, flip angle 90°; 44 slices, voxel size 3.5 × 3.5 × 3.5 mm³) with interleaved slice acquisition.

##### Two-tensor UKF / Free-water Analysis

Diffusion-weighted MRI data were modeled using a two-tensor Unscented Kalman Filter (UKF) framework, which performs recursive estimation of local fiber orientation and diffusion parameters along streamline trajectories and is specifically designed to handle crossing fibers and regions of edema^41,42^. This approach has been validated in both research and clinical settings, including neurosurgical tractography and comparative evaluations of diffusion models on clinical scans^43^. For each voxel, the two-tensor UKF model yielded a multi-compartment representation comprising a free-water compartment and up to two anisotropic tissue compartments^44^. From these compartments, we derived free-water fraction as well as standard diffusion tensor imaging (DTI) indices for fractional anisotropy (FA), axial diffusivity (AD), radial diffusivity (RD), mean diffusivity (MD) and tensor-specific diffusion volumes (or volume fractions). All FA, AD, RD and MD measures reported in this study were obtained from UKF-based fits that explicitly modeled and adjusted for Free-water. Free-water was modeled as an isotropic compartment with diffusivity consistent with freely diffusing water at body temperature, and the free-water fraction was defined as the proportion of the voxel volume attributable to this compartment^45^. The remaining tissue signal was used to estimate free-water–corrected FA, AD, RD and MD, reducing partial-volume contamination from cerebrospinal fluid (CSF) and extracellular fluid and improving sensitivity to microstructural changes within white matter.

##### Free-water groupwise comparisons

Within each network, groups were compared at each FW region using Welch’s two-sample *t*-tests (unequal variances) applied to the FW z-scores. Effects were summarized as the difference in group means, the *t*-statistic, and the two-tailed *p*-value. Because we had directional hypotheses regarding FW differences consistent with cumulative exposure effects, we also report one-tailed *p*-values for the observed direction by halving the two-tailed *p*. No multiple-comparisons correction was applied, so results should be interpreted as hypothesis-generating rather than definitive with the exception for right insula which was evaluated *a priori* based on murine and human lesion patterns.

##### Exposure-Injury Mapping

We investigated the association between cumulative blast exposure using the proxy of time in Low-Level Blast Occupation and brain microstructure as measured by DTI. Diffusion-weighted images were preprocessed using MRtrix3, including denoising, correction for Gibbs ringing, motion correction and eddy-current–related distortions^46^. Preprocessing generated quality-controlled diffusion datasets for each participant as well as summary outputs suitable for group-level analysis. Two-tensor diffusion modeling and tractography were then performed using the UKF tractography implementation in 3D Slicer, yielding voxelwise and tract-derived metrics for each tensor. Detailed preprocessing and tractography scripts are provided in the **Supplementary Information**. For statistical analysis, we computed age-adjusted partial correlations between each DTI-derived metric and Time in Low-Level Blast Occupation, adjusted for age. Partial correlation coefficients and corresponding *p*-values were summarized for each diffusion metric to characterize the relationship between cumulative blast exposure and white-matter injury.

##### Neurite Orientation Dispersion and Density Imaging (NODDI)

Diffusion-weighted images were analyzed using the NODDI_toolbox (version 1.05; MATLAB-based NODDI implementation)^47^. This toolbox fits a multi-compartment diffusion model that decomposes the signal within each voxel into contributions from intra-neurite, extra-neurite and isotropic (free-water) compartments, yielding quantitative maps of neurite density index, orientation dispersion index and isotropic volume fraction.

#### Clinical Behavioral Assessments

Depressive symptoms, subjective well-being and sleep quality were assessed using validated self-report instruments. Depressive symptoms were measured with the 9-item Patient Health Questionnaire (PHQ-9), which assesses DSM-5 depressive symptom criteria over the prior two weeks on a 4-point Likert scale (0-3; total score range 0-27, higher scores indicating more severe depression). The PHQ-9 has demonstrated good reliability and diagnostic validity in individuals with traumatic brain injury and in military and Veteran populations, including Special Operations Forces cohorts^48,49,50,51^.

Subjective well-being was measured with the 14-item Mental Health Continuum-Short Form (MHC-SF), which indexes emotional, psychological and social well-being using 6-point frequency ratings (0-5; higher scores indicating greater positive mental health). The MHC-SF has shown robust performance and measurement invariance in Canadian Armed Forces personnel and in US military Veterans, supporting its use in military and Veteran samples^52,53^.

Sleep quality over the past month was assessed with the Pittsburgh Sleep Quality Index (PSQI), a 19-item instrument yielding a global score from 0 to 21, with higher scores reflecting poorer sleep. Global scores >5 are commonly used to indicate clinically significant sleep disturbance. The PSQI is widely used and well validated in adults with TBI and in military and Veteran cohorts, including those with blast-related mTBI and comorbid PTSD^54,55^.

Post-traumatic stress symptom severity was assessed using the PTSD Checklist-5 total score. The PCL-5 is a Likert-scaled instrument assessing thought intrusion, avoidance, negative alterations in cognition/mood, and arousal/reactivity symptoms over the past month, and produces a total severity score (0–80) validated for use with military populations^56,57^. Higher PCL-5 total scores indicate greater post-traumatic stress.

#### Statistical Analyses

##### Groupwise

Participants were drawn from an operational military cohort with available neuroimaging and symptom data at a single scan time point. For select analyses, personnel were stratified into low-versus high-exposure groups based on binary (zero vs non-zero) time in Low-Level Blast Occupation at scanning. Among those with non-missing values, 57 were classified as low- and 31 as high-exposure. Whenever noted, Time in Low-Level Blast Occupation, PTSD symptom severity (PCL-5 total score) and age were used as covariates and or outcomes of interest.

##### Composite construction

To summarize related tracts values (e.g., right Insula AD tensor 1 and tensor 2), composite metrics were created by summing related tensor measures. If either component of a composite was missing for a given participant, the composite was treated as missing for that participant.

##### Z-score transformation

To place all measures on a common scale and facilitate comparison of effect sizes, all numeric variables (including created composites) were standardized to z-scores. The z-transformation was performed once across the full sample prior to stratification by group. For each z-scored outcome variable, we estimated up to three nested ordinary least squares (OLS) linear models with group membership as the primary predictor of interest: 1. Unadjusted model: Z = β0 + βG · group + ε. 2. Age-adjusted model: Z = β0 + βG · group + βA · Age + ε. 3. Age + PTSD-adjusted model: Z = β0 + βG · group + βA · age + βP · PCL_5_TOT + ε; Here, Z denotes the z-scored outcome, group is coded 0 = new and 1 = experienced, age in years, and PCL_5_TOT is the raw PCL-5 total score. For each model, cases with missing data on any included variable (outcome, group, age, PTSD) were dropped (listwise deletion). *Groupwise Effect sizes.* For each model and outcome, the primary quantity of interest was the regression coefficient βG for group, representing the adjusted standardized mean difference between low- and high-exposure groups. We report βG, its standard error, *t*-statistic, two-sided *p*-value, and an approximate Cohen’s *d*, computed as: *d* ≈ 2 · t / √df_resid; where df_resid is the residual degrees of freedom for the model. This approximation treats the standardized group effect as comparable to a Cohen’s *d* derived from a two-sample *t*-test.

Positive values of βG and d indicate higher z-scores in experienced personnel relative to new personnel. No correction for multiple comparisons was applied at this stage. *Implementation.* All analyses were conducted in either R or Python using pandas for data management and statsmodels for OLS regression.

##### Ordinal (5-band) Bayesian Cumulative Logit

We fit separate ordinal cumulative logit models (5 PHQ-9 bins: minimal 0-4, mild 5-9, moderate 10-14, moderately severe 15-19, severe ≥ 20) for each predictor. Predictors were standardized (z-scored). Models used Normal(0,1) priors on slope with a weak ridge on cut points. Maximum *a posteriori* (MAP) was estimated via Nelder-Mead. The output dose-response curves show the predicted bin probabilities over −2.5 to +2.5 SD of the predictor. Effect size per bin was defined as Δp = P(bin | +1 SD) − P(bin | −1 SD). Uncertainty is estimated as Bayesian Laplace posterior draws (1000 multivariate normal samples around the MAP using the inverse finite-difference Hessian).

##### Bayesian Probability Curves (Moderated Logistic Model)

We fit a logistic model for PHQ-9 ≥10: logit P(Y≥10) = β0 + β1·Xz + β2·Wz + β3·(Xz·Wz) + β4·AGE_z + β5·PCL_z, where W is the moderator candidate standardized to Wz. Posterior draws were obtained via a Laplace normal approximation around the MLE (multivariate normal with the frequentist covariance). We generated posterior median probability curves at Wz specific SDs as noted and at AGE_z=PCL_z=0, plotted against raw months. This directly visualizes adjusted risk trajectories for clinically significant depression.

##### Interventional Multi-Mediator Model (IMM)

We quantified the interventional indirect effect (ΔPé) of low-level blast exposure on the probability of clinically significant depression. The exposure was X = cumulative LLB exposure (Time in Low-Level Blast Occupation in months). The outcome was Y = I[PHQ9 ≥ 10]. Adjusters were Z = {age at scan, PCL5 total score}. Mediators were selected on Principal Component Analysis loadings from PCA1, which accounted for 36% of variance and focused on features of diffuse insula-basal ganglia circuit injury. Selected mediators were right insula ODI; right insula Free-water; the composite score of right globus pallidus FA tensor1 and tensor2; and the right caudate FA tensor1 and tensor2 composite. We did not use putamen or other measures in order to minimize model complexity and overfitting. Composites were used to preserve conservative assessments of the two tensor UKF method.

Missing mediator values were mean-imputed within-sample to preserve statistical power, with the expectation that this approach biases estimates conservatively toward the null. A-paths (X → M): Each mediator M_j was modeled via ordinary least squares: M_j = α0j + αXj·X + αZj^T·Z + ε_j. We report β_a = αXj with 95% CIs from the OLS covariance. B- and cʹ-paths (M → Y, X → Y): The binary outcome was modeled with logistic regression: logit P(Y=1) = β0 + βX·X + Σ_j βMj·M_j + βZ^T·Z. We report β_bj = βMj and cʹ = βX with 95% CIs from the observed Fisher information.

##### IMM-Moderator Outcome Modeling

Moderator effects were evaluated as a function of IMM vs exposure. A-paths (OLS) were calculated via M = α0 + αX·X + αAGE·AGE + αPCL·PCL + ε. Outcomes were modeled using logistic, ridge-regularized IRLS as a logistic probability of PHQ9 ≥ 10 using logit P(Y=1) = β0 + βX·X + βM·M + βW·W + βMW·(M×W) + βAGE·AGE + βPCL·PCL. We draw from multivariate normal approximations to the a-path OLS and outcome-model IRLS covariance matrices (2,500 iterations) to produce posterior predictive distributions of P(Y=1) along an exposure grid in raw months with AGE_z = PCL_z = 0. The moderator W is fixed at specified SD levels to generate Low and High curves with 95% credible bands. *Dual Moderator-Risk Outcome Modeling.* We quantified how combinations of two moderators (W1, W2) modify the interventional effect of low-level blast exposure on depression risk. Exposure was X = Time in Low-Level Blast Occupation. The binary outcome was Y = I[PHQ-9 total score ≥ 10]. Analyses used complete cases on X, Y, age, and PCL5 total scores (N = 67). All models were adjusted for age and PTSD symptom severity (PCL5). In surface calculations, age and PCL were fixed at their sample means (thus effects are shown at average covariate levels).

##### Mediator

The four candidate mediators were used: (1) can_ctx_rh_insula_ODI; (2) INSULAR_right_Free-water (z-scored within-sample); (3) GlobusPal_R_FA_comp = z(mean of 150_GlobusPal_R_FA_tensor1, 150_GlobusPal_R_FA_tensor2); (4) Caudate_R_FA_comp = z(mean of 147_Caudate_R_FA_tensor1, 147_Caudate_R_FA_tensor2). Missing mediator values were mean-imputed within-sample prior to z-scoring for the Free-water/FA composites. <u>A-paths</u> <u>(X → M).</u> For each mediator M_j we fit ordinary least squares with adjustment: M_j = α0j + αXj·X + αAGEj·AGE + αPCLj·PCL + ε_j. These α^ coefficients are used to compute the expected mediator levels when X is set to +2 SD versus 0 (holding age, PCL5 at their means).

##### Outcome model (moderation with two moderators)

For each ordered pair (W1, W2), we fit a penalized logistic regression for Y with the following specification: logit P(Y=1) = β0 + βX·X + Σ_j βMj·M_j + βW1·W1 + βW2·W2 + βW1W2·(W1×W2) + Σ_j [ β(Mj×W1)·(M_j×W1) + β(Mj×W2)·(M_j×W2)] + βAGE·AGE + βPCL·PCL. Moderators were z-standardized (within-sample) with mean-imputation for missing values. Ridge (L2) regularization (λ = 0.8) stabilized estimation in the multicollinear, small-N method. <u>ΔP-from-M surface</u>. For each (W1, W2) grid point on [−3, +3]×[−3, +3] (41×41), we computed ΔP = E_Z [ P(Y=1 | X=+2, M(X=+2), W1, W2, Zé) − P(Y=1 | X=0, M(X=0), W1, W2, Zé)], where M(X=+2) and M(X=0) are obtained from the A-path OLS equations, and Zé denotes age/PCL5 fixed at their sample means. Each W1 run yields 18 panels (one per W2 ≠ W1).

## Results

### Highly repetitive, 3 psi Low-Level Blast Causes Chronic White Matter Injury in the Insula, Basal Ganglia and associated Subcortical Forebrain Structures in Mice

Mice received 6 daily blasts of 3 psi (Monday through Friday) for 3 consecutive weeks; sham controls received equal anesthesia without blast exposure^24^. After the final exposure, mice recovered for six months before high-resolution gadolinium-enhanced 14T *ex vivo* diffusion MRI (**Fig. 1a**). Blast overpressure characteristics from 80 randomly selected pressure profiles were 3.27 ± 0.11 peak psi, 1.2 ± 0.02 ms positive-phase duration, and impulse of 3.94 ± 0.132 psi·ms (mean ± s.e.m.; **Fig. 1b**).

**Figure 1.**
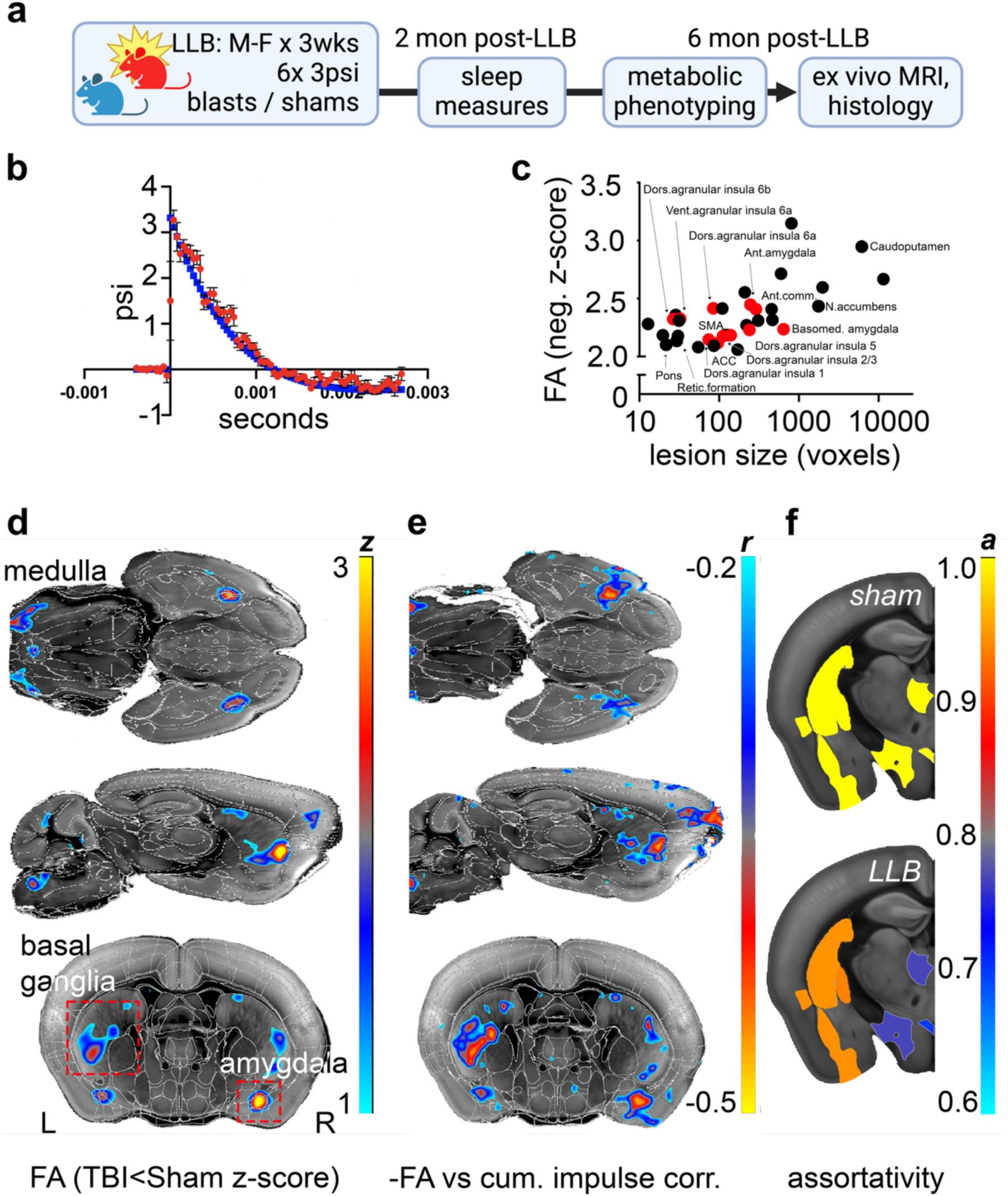
Low-level blast (LLB) exposure causes chronic white matter injury in the insular Central Autonomic Network and corpus striatum. a, Mouse procedural timeline. Mice were exposed to 6 LLB blasts per day measuring ∼3 psi per blast. LLB occurred Monday through Friday for 3 consecutive weeks (90 total LLB blasts) before recovering without medical intervention in their home cage for 6 months before undergoing ex vivo 14T diffusion tensor imaging. Sham control mice experienced equal handling but did not receive any blast exposures. b, Average blast overpressure vs. time curve (red) for 80 randomly-selected study blast waves compared with the idealized open-field Friedlander wave (blue). c, Brain regional z-scores vs. number of regional voxels with significantly reduced fractional anisotropy (FA) in LLB mice compared to sham control mouse values on *ex vivo* 14T dMRI. Red dots are areas of both the Central Autonomic Network and the default Ascending Arousal Network. d, *z*-score map for (c). e, Correlation map (*r*, *r*-values) of FA vs. cumulative impulse. f, LLB reduces relative measures of connectivity (assortativity; *a*-values) in salience network components insula, caudatoputamen, globus pallidus, lateral hypothalamic area, mediodorsal nucleus of thalamus, and paraventricular hypothalamic nucleus as compared to values in sham control mice. N=7 sham, 12 LLB mice.

Whole-brain analyses aligned to the Allen Common Coordinate Framework^36^ demonstrated significant reductions in diffusion-derived fractional anisotropy (FA) across multiple forebrain regions, consistent with chronic microstructural compromise (**Fig. 1c,d**). Injured areas were enriched in an insula-centered circuit, including agranular insular subfields (input and output layers), insular-associated white matter (external capsule), claustrum, caudoputamen, nucleus accumbens, and anterior commissure, with additional involvement in amygdala- and brainstem-adjacent areas (**Fig. 1c,d**). Within these regions, FA reductions were correlated with cumulative blast impulse (**Fig. 1e**), linking dosimetry to chronic microstructural injury.

To assess circuit organization, we quantified structural connectivity assortativity (**Fig. 1f**). Positive assortativity was reduced by LLB in key nodes of the same circuit (N=7 sham, 12 LLB; two-tailed tests): agranular dorsal insula layer 6a (*t*[1]=2.16; *p*=0.045), caudoputamen (*t*[1]=2.32; *p*=0.033), dorsal pallidum including globus pallidus (*t*[1]=2.57; *p*=0.020), paraventricular hypothalamic nucleus (*t*[1]=3.24; *p*=0.005), mediodorsal thalamic nucleus (*t*[1]=2.66; *p*=0.016), lateral hypothalamus (*t*[1]=3.26; *p*=0.0046), and caudal pallidum with substantia innominata (*t*[1]=2.64; *p*=0.017). Similar trends were observed in ventral pallidum but did not reach significance. Together, these results indicate that highly repetitive sub-threshold blast exposure produces long-lasting white-matter injury and reduced microstructural network organization in an insula-basal ganglia-thalamic circuit.

### Experimental Highly Repetitive Low-Level Blast (3 psi) Causes Chronic Insular and Subcortical Forebrain Neurodegeneration in Mice

To determine whether diffusion and network changes correspond to chronic neuropathology, we examined neuroimmune and neurodegenerative markers in brains from the diffusion cohort (**Fig. 2**). Compared with sham tissues, LLB induced diffuse GFAP-positive astrogliosis across forebrain structures, with prominent increases along lateral cortex, ventricular boundaries, and myelinated insertions traversing caudoputamen. IBA1^+^ microglia/monocytes were also diffuse, whereas CD68^+^ macrophages showed focal enrichment in the basal forebrain and colocalized with AT8^+^ tauopathy. pTau217 expression was highest in insula and basal forebrain (**Fig. 2a-d**), and GFAP expression followed the topography of insular pTau217 without clear interface scarring, consistent with chronic but subtle glial remodeling (**Fig. 2e,f**).

**Figure 2.**
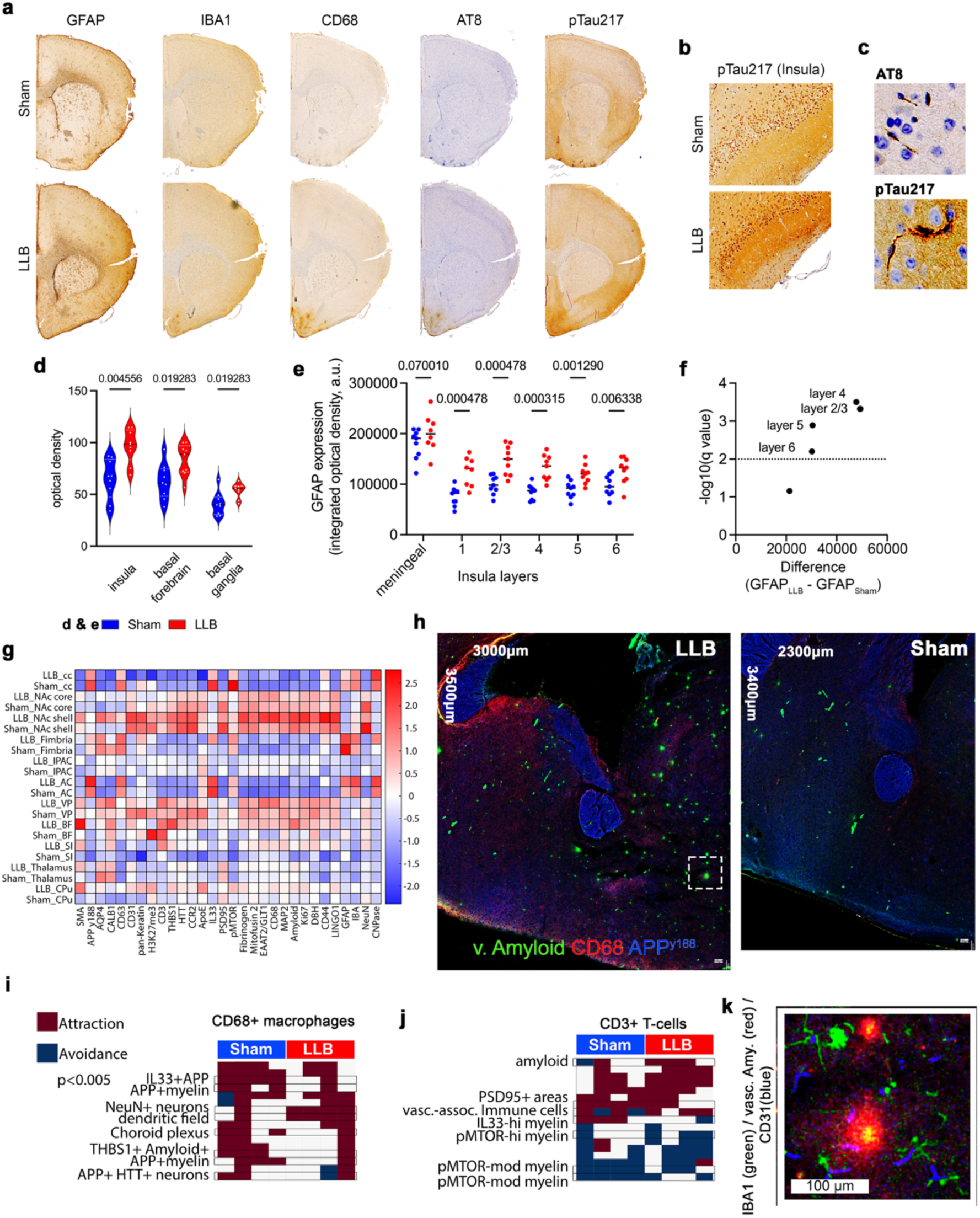
Low-level blast (LLB) induces chronic basal forebrain neurodegeneration and tauopathy in insula and subcortical forebrain in mice. **a,** Example neuropathological series from a single LLB and yoked sham mouse from Figure 1. LLB-induced changes in forebrain GFAP and IBA1 are relatively subtle. However, macrophage marker CD68 and early neurofibrillary tangle and neuropil thread marker AT8 is apparent in the basal forebrain. **b,** pTau217-stained insula from a yoked pair of study mice. All tissue sections were stained on slides with a both a unique sham and LLB mouse represented. **c)** Higher magnification insets with representative AT8 and pTau217-positive staining. Using LLB mice, AT8 was imaged from basal forebrain; pTau217 was imaged from insula. **d,** Quantitation of pTau217 optical densities of all study mice across entire anatomical regions for insula, basal forebrain and basal ganglia. pTau217 is an accurate and sensitive biomarker for detecting the early accumulation of both amyloid-beta (Aβ) plaques and tau pathology in Alzheimer’s disease (AD) than AT8. **e)** GFAP quantitation across insula layers. **f,** 1% False Discovery Rate-adjusted discovery analysis results of GFAP quantitation in insula layers. **g)** Mean imaging mass cytometry (IMC) marker expression across forebrain structures in 4 sham vs. 5 LLB mouse brains. LLB increases vascular amyloid expression in the basal forebrain, thalamus and caudoputamen and aquaporin-4 amongst most striatal regions. **h,** Representative IMC images from LLB (left) and sham mice (right) showing regional distribution and intensity of vascular amyloid (green) and CD68 expression (red). **I,** Neighborhood analysis of LLB-induced changes in cellular associations of CD68+ microglia and **(i)** CD3+ T-cells. LLB reduces macrophage associations with Thbs1^+^/vascular amyloid^+^ and APP^y188+^ structures while increasing associations between CD3+ cells with amyloid and vascular immune cells. **k,** LLB mouse from **(h, box)** showing vascular amyloid appears as parenchymal deposits occurring outside of CD31^+^ (or SMA^+^; data not shown) vasculature accompanied by minimal GFAP^+^ reactive astrocytosis and little-to-mild microgliopathy. **d,e,** Dots represent results from individual mice. N=9 per group. **g-j,** N=4 sham, 5 LLB mice, 6 month recovery time point.

We then profiled ventral forebrain pathology using imaging mass cytometry (IMC) with an established 35-antibody panel capturing structural, signaling, vascular, and immune markers of murine TBI pathology (4 sham, 5 LLB; **Fig. 2g-k**)^58^. Across basal regions, LLB increased smooth muscle actin (SMA), consistent with vascular-associated remodeling. Aquaporin-4 (AQP4) showed region-dependent changes, increasing in caudoputamen, ventral pallidum, and nucleus accumbens shell/core and decreasing in thalamus, implicating altered water regulation during chronic recovery^59,60,61^. Consistent with this, *aqp*4 mRNA levels in forebrain preparations were increased by LLB (*t(*45.41)=3.095, *p*=0.0034; **Supplemental data**). CCR2^+^ peripheral immune cells were most evident in caudoputamen and basal forebrain, and CD68 levels were increased across basal structures with the largest changes in basal forebrain, substantia innominata, and ventral pallidum.

IMC additionally indicated vascular-adjacent amyloid accumulation across caudoputamen, substantia innominata, basal forebrain, nucleus accumbens shell, and thalamus. Neighborhood analyses suggested that LLB reduced global interactions of CD68^+^ macrophages with IL33^+^, APP^y188+^, and vascular amyloid^+^ cells, shifting macrophage associations toward NeuN^+^ neurons and dendritic/axonal fields, while increasing CD3^+^ cellular attractions to amyloid^+^ areas and vascular-associated immune cells (**Fig. 2h-k**). Collectively, these data support a chronic injury state after repetitive sub-threshold blast characterized by insula/basal forebrain tauopathy, glial and macrophage activation, and peri-vascular amyloid and adjacent remodeling that aligns with the microstructural network signature.

### LLB Exposure Causes Sleep and Metabolic Deficits in Mice Correlated with Insula and Basal Ganglia Assortative Loss

Sleep disturbance is commonly endorsed after blast exposure^62^ and can potentiate mood dysregulation^63^. Two months after exposure, LLB mice showed increased daily sleep percentage and increased total sleep bout length, with a trend toward increased diurnal wake ratio across a three-day recording period (**Fig. 3a**). Mean bout length linearly scaled with cumulative blast impulse (*F*(1,18)=5.20, *p*=0.035) and inversely related to six-month caudoputamen assortativity (*F*(1,17)=6.18, *p*=0.024), linking early sleep disruption to chronic network-level compromise.

**Figure 3.**
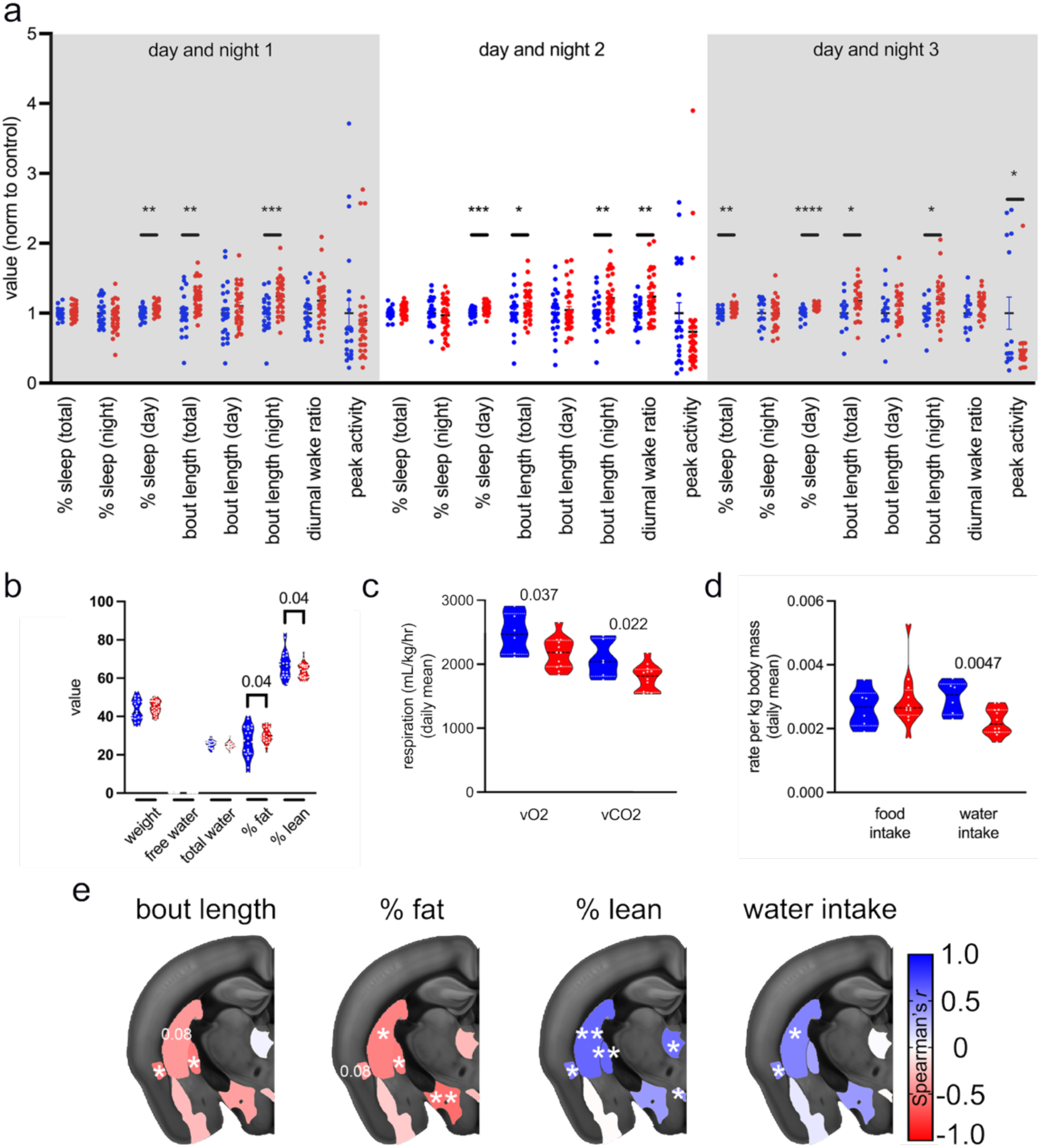
LLB induces chronic sleep and metabolic injuries, behaviors regulated by the basal forebrain. **a,** Graph represents parameterized sleep results from individual LLB (red) and sham mice (blue)(dots) measured over 3 consecutive days, 2-months after the final LLB or sham exposure. LLB increases sleep bout length and daytime sleepiness. **b,** Body composition measures 6-months after LLB and sham exposures. LLB exposure reduces lean body mass and increase percent body fat. **c)** Respiratory rate for vO2 and vCO2 is decreased in LLB mice. **d,** Daily food and water intakes. Water intake is decreased in LLB-exposed mice. Dots represent results from individual animals. **e,** Spearman’s *r*-mapping for significant region-behavior correlations. \**p*<0.05, \*\**p*<0.01, \*\*\**p*<0.001. Sample sizes are **(a)** N=23 sham, 32 LLB; **(b-e)** N=6 Sham, 12 LLB, respectively. **a-d,** 2-tailed t-tests. Red, LLB; blue, sham.

We next assessed body composition and metabolism at six months. Total body weight and total/Free-water measures were unchanged, but percent fat increased (*t*[50]=2.068; *p*=0.044) and percent lean mass decreased proportionally (*t*[50]=2.07; *p*=0.044)(**Fig. 3b**). Indirect calorimetry revealed lower oxygen consumption and carbon dioxide production in LLB mice (vO2 *p*=0.037; vCO2 *p*=0.022), with the respiratory exchange ratio trending lower (*p*=0.076), consistent with altered substrate utilization (**Fig. 3c**). Food intake did not differ, but water consumption per gram of body weight was reduced (*p*=0.0047; **Fig. 3d**). These altered behavioral and metabolic measures correlated with assortativity across insula, caudoputamen, and dorsal pallidum, and assortativity measures predicted water intake and adiposity measures in regression models (**Fig. 3e**). Together, these findings indicate that repetitive LLB produces persistent physiologic alterations that track disruption of an insula-basal ganglia circuit implicated in sleep, autonomic regulation, and energy balance.

### Worse Insular Central Autonomic Network Injury is Associated with Increased Career LLB Exposure in US Special Operations Forces Soldiers

To test whether the insula-centered injury signature extends to an operationally-relevant human cohort, we conducted an age-adjusted voxel-wise diffusion MRI analysis in newly-versus previously-certified, in-career US Army SOF Soldiers representing Low- and High-career LLB exposure groups relative to one another (N= 27 New/Low-exposure, 28 Experienced/High-exposure SOF Soldiers). We used Time in Low-Level Blast Occupation (i.e., months from certification to acquisition of study measures) as a previously defined and empirically supported proxy of cumulative LLB burden in this cohort^29,64^. Groups had similar demographics and service histories (**Supplemental Table 1**). SOF soldiers completing their organization’s training pipeline were aged 32.3 ± 0.7 years with 11.4 ± 0.7 years of combat arms military service before completion (ranges, age: 24-38 years, prior service: 6-16 years; 86.7% as combat arms military occupational specialties before SOF career); in-career Experienced SOF were aged 35.3 ± 0.7 years with 10.8 ± 0.6 years military service before their SOF career and 4.9 ± 0.4 years of SOF career length post-certification (ranges, age: 27-45 years; SOF career length: 0.8-10.6 years; 85.7% as combat arms military occupational specialties). Unbiased, age-adjusted, voxel-wise analyses identified a large, contiguous region of reduced FA around the dorsal, medial, and ventral perimeter of the right insula that correlated with cumulative LLB burden (**Fig. 4a**).

**Figure 4.**
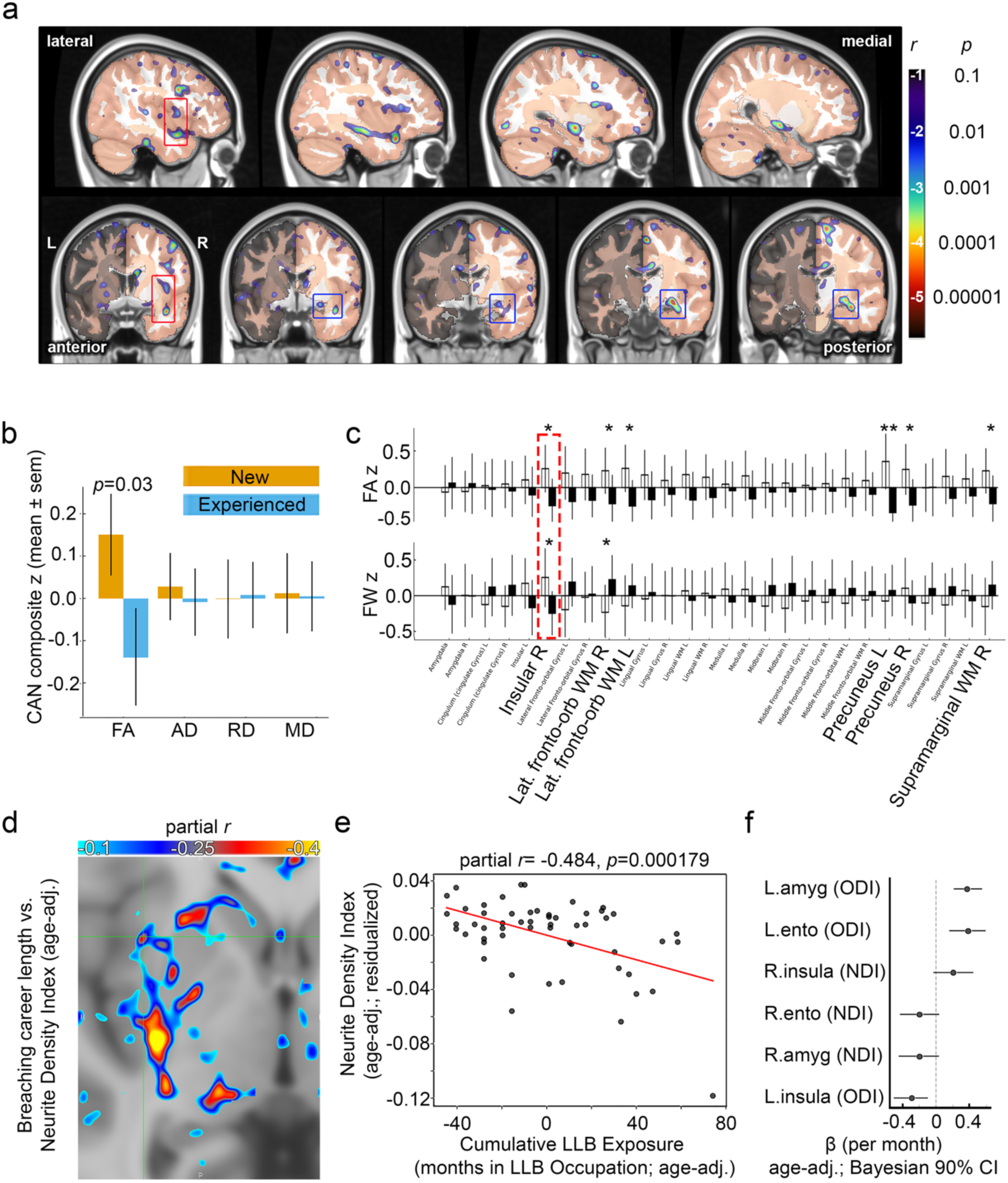
In military Service members, right insula-centered diffusion abnormalities are associated with cumulative low-level blast exposure. **a,** Voxelwise brainwide maps of age-adjusted partial Pearson correlation coefficients between cumulative low-level blast exposure (Time in LLB Occupation, months) and free-water-corrected fractional anisotropy (FA). Colors encode the strength and direction of the association (color bar, partial *r* with corresponding *p* values). A prominent contiguous cluster of reduced FA is localized to the right insula and adjacent connected structures. **b,** Central autonomic network (CAN) composite z-scores for FA, axial diffusivity (AD), radial diffusivity (RD) and mean diffusivity (MD) in New (certified SOF with 0 months in LLB Occupation) versus Experienced (certified SOF with >0 months) SOF Soldiers. Bars show mean ± s.e.m.; CAN FA is significantly lower in Experienced SOF Soldiers (*p* = 0.03), whereas other CAN diffusion composites and salience/SOFA network composites (not shown) are not significantly different. **c,** Region-wise CAN profiles for FA (top) and Free-water (FW; bottom), plotted as z-scored group means ± s.e.m. across bilateral CAN regions. Asterisks denote group differences (*p* < 0.05; \**p* < 0.01). The red dashed box highlights reduced FA and FW in the right insula. **d,** Voxelwise lesion map of age-adjusted partial correlations between Time in LLB Occupation and neurite density index (NDI). Significant negative associations (cool colors) are observed in right insula, caudate, amygdala and entorhinal cortex. **e,** Scatterplot of residualized right insula NDI from lesions versus cumulative LLB exposure (age-adjusted Time in LLB Occupation; months). Each dot is a participant; the red line shows the regression fit (partial *r* = −0.484, *p* = 0.000179), indicating lower neurite density with greater SOF Time in LLB Occupation. **f,** Bayesian whole-region estimates of the effect of SOF Time in LLB Occupation on NDI and orientation dispersion index (ODI) in bilateral amygdala, entorhinal cortex and insula. Points represent posterior median β (per month, age-adjusted) and horizontal lines the 90% credible intervals; the vertical dashed line indicates no effect. Negative values reflect progressive reductions in neurite density or complexity with greater cumulative exposure. All analyses in **a, c-f** use Pearson’s partial correlations controlling for age; N=55 participants.

We then evaluated diffusion composites derived from *a priori* network definitions and supported by the observed injury patterns in LLB mice. Mean FA scores were lower in the insula-hubbed central autonomic network (CAN) in experienced SOF compared with new SOF under one-tailed testing (*p*=0.03; **Fig. 4b**). Comparable composite analyses for Salience and Striatal-Orbitofrontal-Amygdala (SOFA) networks did not reach significance across diffusion measures, indicating that exposure-linked microstructural vulnerability is enriched among CAN regions. Exploratory region-level analyses of group-wise FA and Free-water (FW) differences highlighted several CAN structures, including the right insula, with age-adjusted differences in FA and FW consistent with microstructural injury (each ∼*t*[65]=2.0; FA *p*=0.05, FW *p*=0.05, 2-tailed; **Fig. 4c**).

To further characterize microstructure, we assessed neurite orientation dispersion and density imaging (NODDI). Age-adjusted neurite density index (NDI) was negatively correlated with SOF career length within lesion-like areas of insula, caudate, amygdala, and entorhinal cortex (**Fig. 4d,e**), while overall NDI increased in the right insula (**Fig. 4f**), suggesting heterogeneous microstructural responses within the insula that may reflect combined changes in neurite packing and numbers, extracellular water, and fiber geometry. Orientation dispersion index (ODI) decreased with cumulative exposure, with a significant reduction in left insula ODI and strong posterior support for decreased right insula ODI in Bayesian analyses (left insula ODI: [β=-0.2993, [95% CI, −0.5403 to −0.0583], *p*=0.0157; P<0=0.992]; right insula ODI: [β=-0.1485, [95% CI, −0.3906 to 0.0936], *p*=0.225; P<0=0.888]; **Supplemental Data**). Together these results indicate that cumulative LLB exposure in SOF soldiers is associated with diffuse insular and central autonomic network microstructural injury that mirrors exposure-associated injury patterns observed in our empirically parameterized mouse model.

### Emotional and Psychological Endorsements Worsen with Longer SOF Career Length

The insula integrates broad informational streams as emotional outputs for downstream executive functions^65^ which we hypothesized are skewed towards more frequent endorsements of depression and anxiety in persons with a history of blast-induced neurotrauma. We next assessed depression, wellbeing, and sleep in 146 SOF soldiers and pre-certified SOF trainees (**Supplemental Table 1**). PHQ-9 scores were higher in experienced in-career SOF Soldiers and retired SOF Veterans compared to trainees (N=31 trainees, N=51 experienced in-career, N=10 retirees; means ± s.e.m.: 2.29 ± 0.49, 6.67 ± 0.65, 6.00 ± 1.36; trainee vs. in-career *p*<0.0001; trainee vs. retiree *p*=0.0265; **Fig. 5a**).

**Figure 5.**
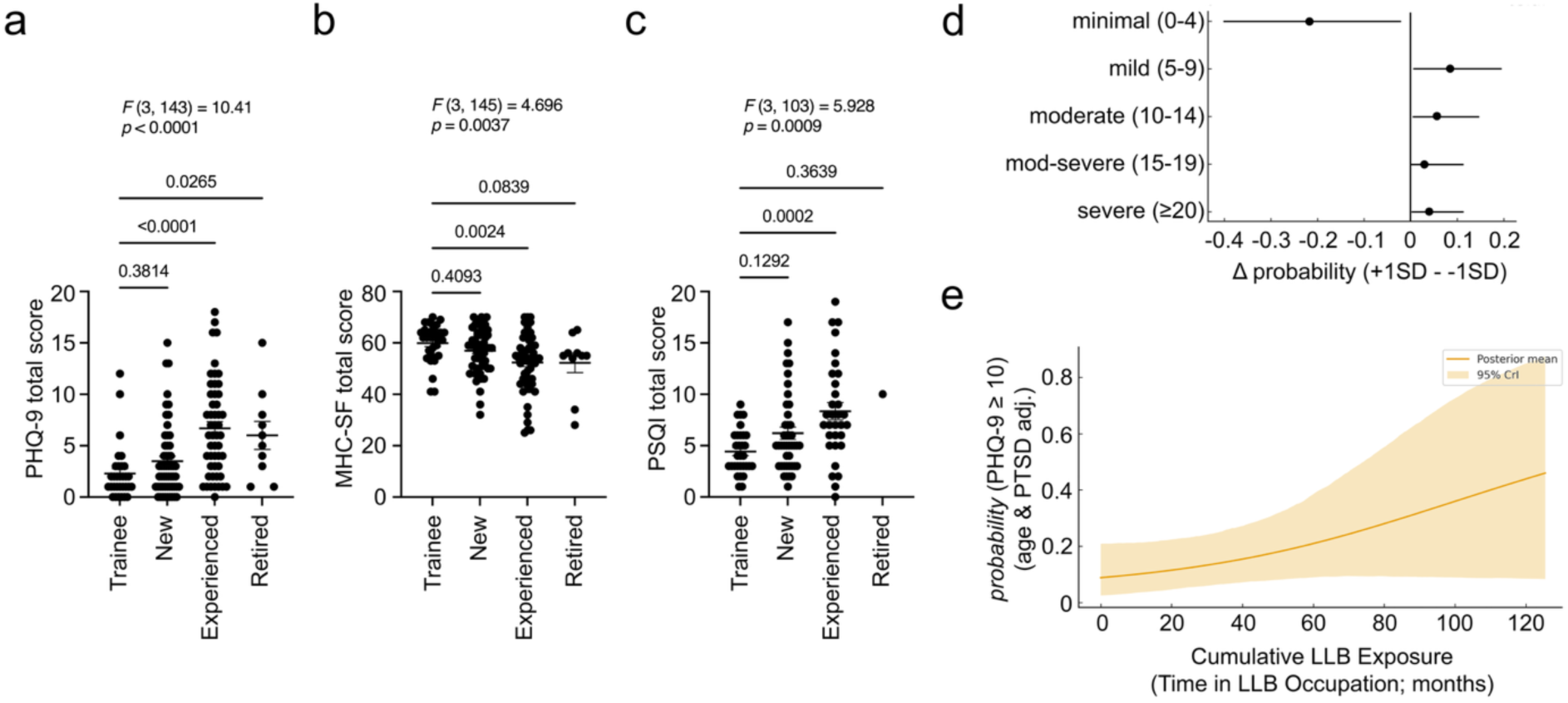
The Probability of Clinically Significant Depression Increases with Career Low-Level Blast Exposure. Total scores across career stages in US Army Special Operations Forces Soldiers for **a,** Patient Health Questionnaire-9 (depression severity), **b,** Mental Health Continuum-Short Form (Psychosocial emotional wellbeing), **c,** Pittsburgh Sleep Quality Index (Sleep Injury) by career stage. **d,** Ordinal (5-band) Bayesian Cumulative Logit with per-bin probabilities based on PHQ-9 cutoffs for depression severity categories. Entries are cross-validated mean predicted probabilities for each PHQ-9 category. In age- and PTSD-adjusted models, a +1 SD change SOF career length increases the combined probability of PHQ-9 ≥10 by 0.15. Largest shift in the mild bin. **e,** Age- and PTSD-adjusted Bayesian posterior mean probability (solid) with 95% credible interval (shaded), x-axis in raw months (mean 25.4, SD 33.3). **a-c,** Dots represent results from individual participants. N=31 SOF trainees, 43-55 New SOF 32-51 Experienced SOF Breachers, 1-10 Retired SOF. 2-way ANOVA with 2-tailed post hoc *t*-tests.

Positive mental health was assessed using the MHC-SF^52,53^. Scores worsened with later career stage (*p*=0.0042). Total MHC-SF was statistically indistinguishable between SOF trainees and newly-certified SOF soldiers, but was significantly lower in experienced in-career SOF Soldiers compared to trainees or New SOF (*p*=0.0032; **Fig. 5b**), indicating reduced wellbeing in emotional, psychological, and social domains. Using multivariate linear regression controlling for group and age (excluding retirees due to missing SOF career length), SOF career stage was significantly predictive of worsening scores for MHC-SF total (*p*=0.0037).

Sleep quality was assessed using the Pittsburgh Sleep Quality Index (PSQI)^54,66,67^. PSQI scores increased across career stages (*p*=0.0009), and experienced SOF Soldiers exceeded new SOF soldiers by approximately four points on average (*p*=0.0002; **Fig. 5c**), consistent with clinically meaningful sleep deterioration^68^.

Using an ordinal (5-band) Bayesian cumulative logit approach with per-bin probabilities based on PHQ-9 severity cutoffs, the probability of moving from a minimal depression status to an increased depression category was significantly increased across SOF careers stages increasing risk of development of clinical depression 15%/time-in-LLB-Occupation SD (**Fig. 5d**); with overall risk of developing clinical depression (PHQ-9 ≥10) estimated to increase ∼25% (range ∼10-50%) per 6-years in LLB occupation in age- and PTSD-adjusted models (**Fig. 5e**). Together these results indicate cumulative LLB burden increases depression risk independent of age and PTSD.

### Diffuse Insula Network Injury Mediates LLB-Induced Depression with Clinical Heterogeneity Moderated by Network Features in US Army SOF Soldiers

Interventional Multi Mediation modeling was used to test whether diffuse right insula-basal ganglia injury could mediate the effect of cumulative LLB exposure (proxied by time in LLB Occupation) to increase depression risk (PHQ-9 ≥10). Principal component analysis loadings for right insula ODI and Free-water, and separate right globus pallidus and right caudate FA composites were used to estimate diffuse neural injury in the efferent pathways linking insula-hubbed cortical streams with downstream executive function. Using this composite as our mediator, the causal mediation estimate of cumulative LLB exposure to increase risk for clinical depression (probability(PHQ9≥10); **Fig. 6a**) was significant. The IMM model estimates a ΔPé =0.243 [0.014, 0.578] with a probability of increasing PHQ-9 scores of 0.984. A-path estimates showed the expected positive associations between SOF time in LLB Occupation and selected microstructural mediators; b-paths indicated that higher mediator values were associated with increased probability of PHQ-9 ≥10 after adjustment for age and PTSD. Modeling the probability of clinical depression risk versus cumulative LLB exposure on the IMM model recapitulated earlier Bayesian results (**Fig. 6b**; **Fig. 5e**).

**Figure 6.**
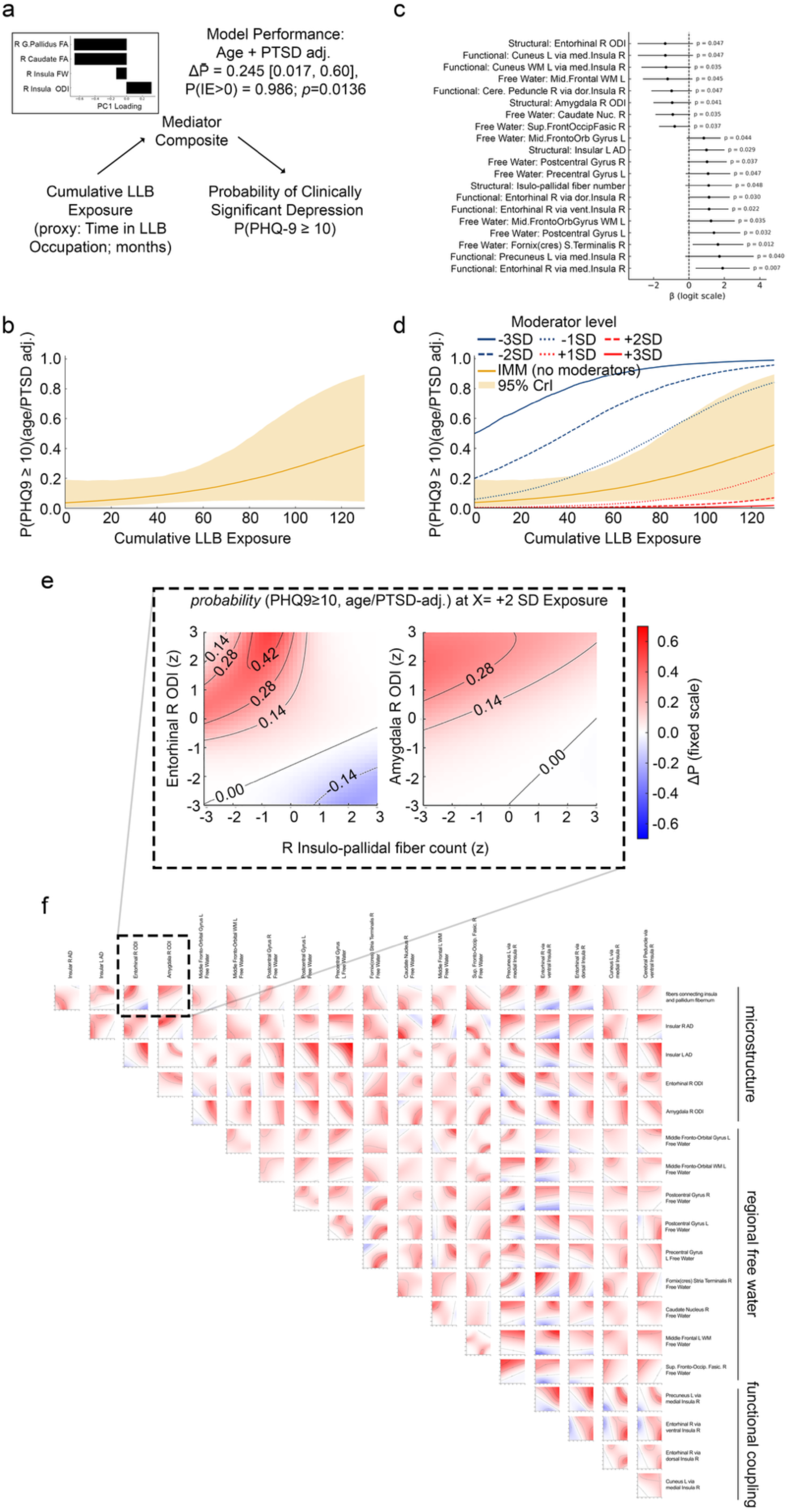
Insula network features moderate heterogeneity in LLB-depression risk. **a,** Principal component analysis of an insula-striatal network (right globus pallidus FA, right caudate FA, right insula Free-water and ODI) was used to derive a PC1-weighted Interventional Multivariate Mediator (IMM) composite. In an age- and PTSD-adjusted model, cumulative LLB exposure (Time in LLB Occupation) increased the probability of PHQ-9 ≥10 through this IMM, with an indirect-effect probability change ΔP = 0.245 (95% credible interval (CrI) 0.017–0.60), posterior probability P(IE>0) = 0.986 and p = 0.0136. **b,** Predicted probability of PHQ-9 ≥10 as a function of cumulative LLB exposure for the IMM in the absence of moderators. Solid line, posterior median; shaded ribbon, 95% CrI. **c,** Forest plot of candidate moderators identified in an unbiased screen. Points indicate age- and PTSD-adjusted logit coefficients (β) for the effect of each candidate on depression risk within the IMM; horizontal bars denote 95% CrIs. Candidates span diffusion microstructure, regional Free-water and task-free fMRI coupling between insula and cortical-subcortical targets. **d,** Example of a single significant moderator, right insulo-pallidal fiber number. Colored curves show the IMM-predicted probability of PHQ-9 ≥10 across cumulative LLB exposure when the moderator is fixed at −3 to +3 SD (legend). The beige band shows the 95% CrI for the IMM without moderators. **e,** Representative two-dimensional moderator surfaces derived at high cumulative LLB exposure (X = +2 SD). Heat maps show the median change in probability of PHQ-9 ≥10 (ΔP; fixed color scale) as a function of right insulo-pallidal fiber count (x axis) and either right entorhinal ODI (left) or right amygdala ODI (right). Warm colors indicate increased risk; cool colors indicate reduced risk (resilience). **f,** Matrix of pairwise moderator maps for all candidate combinations, grouped by modality (microstructure, regional Free-water and functional coupling). Each panel depicts ΔP for PHQ-9 ≥10 at X = +2 SD exposure as a function of the two moderators (both in z units), with red indicating higher and blue indicating lower predicted risk. All estimates are adjusted for age and PTSD symptom severity. N=67 participants.

The IMM model provides strong evidence that cumulative LLB exposure over a SOF career is associated with an increased risk probability of PHQ-9-defined clinically significant depression, consistent with a mediation pathway through diffuse insula-basal ganglia injury. It does not, however, explain the heterogeneity observed in exposure responses. To provide a logical framework for how and why two individuals with similar injury severity among mediator components can experience the same blast loading yet exhibit different levels of depression, we investigated a role for network features to moderate injury-outcome relationships. We used an unbiased, high-dimensional screening approach to identify candidate moderators of the injury-depression relationship. Nineteen moderator candidates with significantly predictive Bayesian posterior probabilities were identified (**Fig. 6c**). Importantly, these moderator effects are not intended to support individual-level prediction, but rather to explore why group-level exposure-outcome relationships may appear inconsistent across individuals.

We then quantified how mediator pathways amplify or mitigate the probability of clinically significant depression (PHQ-9 ≥10) at a fixed high exposure (X = time in Low-Level Blast Occupation = +2 SD) as a function of pairs of moderators (W1, W2), highlighting heterogeneity in risk for the same nominal exposure. Across unique moderator combinations, ΔP-from-mediator surfaces showed non-uniform patterns with distinct risk-increasing and risk-decreasing zones.

Zones often aligned with circuits involving insular microstructure and basal ganglia FA composites, fronto-parietal Free-water, and entorhinal/dorsal streams. Several moderators produced strong positive ΔP regions when both W1 and W2 were high (≥+1-2 SD), consistent with an amplified-risk phenotype, whereas low W1/W2 bins (≤−1-2 SD) frequently exhibited negative ΔP, consistent with resilience despite the same exposure level. The presence of sizable M×W1×W2 contributions indicates that mediator effects are not uniform across individuals and that moderator configurations can flip the sign of ΔP, providing a quantitative explanation for divergent clinical trajectories.

## Discussion

We tested whether repetitive low-level blast exposure at 3 psi, which is below the current 4 psi military safety threshold, is associated with findings that explain the elevated risk of clinical depression observed in U.S. Army SOF Soldiers. Across species, we identify a conserved injury pattern in which empirically parameterized, routine LLB preferentially injures an insula-centered cortico-striato-thalamic circuit. In SOF soldiers, diffuse microstructural compromise of this right insula-basal ganglia circuit mediates the relationship between cumulative occupational exposure and PHQ-9-defined clinical depression risk.

A key strength of this study is the empirical anchoring of exposure. Monitoring data from breaching and close-quarters battle training demonstrate that routine charges under 4 psi can accumulate to substantial weekly and career-long dose characterized by high repetition and cumulative impulse^64^. Our 3 psi, 90-blast mouse paradigm, based on these military training data, therefore provides an operationally grounded representation of repetitive sub-threshold loading rather than a traditional “blast-TBI” model. Likewise, treating time in Low-Level Blast Occupation as the primary exposure metric in our human cohort is consistent with an empirically supported framework in which training frequency and cumulative impulse scale with time in role^29,64^.

Repetitive 3 psi LLB in mice produced a durable, network-level injury signature months after exposure. High-field diffusion MRI revealed reduced FA and disrupted network organization in an insula-striatal-pallidal-thalamic loop implicated in salience processing^69,70^, motivated behavior^69^, affect regulation^69^, and autonomic control^70^. Experimental findings from mice provide mechanistic evidence for this circuit injury, with insular and basal forebrain tau-associated pathology, focal macrophage enrichment, diffuse glial activation, and evidence of vascular-adjacent remodeling and altered water regulation. Although the causal ordering among tau pathology, immune activation, and vascular remodeling cannot be established from these data alone, their convergence on basal forebrain and insula supports a chronic, circuit-relevant injury state after repetitive sub-threshold exposure.

Functionally, mice exhibited sleep and metabolic phenotypes that tracked the degree of network disruption. Increased sleep time and longer sleep bouts^71,72^, increased adiposity with reduced lean mass^73^, reduced metabolic rate^73^, and reduced water intake collectively indicate altered autonomic and energy-balance regulation^74,75^ and these phenotypes correlated with insula/striatal/pallidal network organization measures. These data support a model in which repetitive LLB destabilizes an insula-centered control system that integrates interoceptive signals and autonomic state, thereby shaping downstream executive, affective and motivational function.

In SOF Soldiers, diffusion imaging identified a right insula-centered cluster of reduced FA scaling with time in Low-Level Blast Occupation distinct from the effects of age, and CAN composites were selectively affected relative to Salience and SOFA composites. NODDI findings pointed to exposure-linked microstructural alterations across insula, caudate, amygdala, and entorhinal cortex, consistent with disruption of a fronto-limbic-striatal circuit. Clinically, longer career length was associated with higher PHQ-9 scores, lower wellbeing, and worse sleep, even after adjustment for age and PTSD symptoms. The IMM analysis then connected these levels of evidence by showing that this composite significantly mediated the relationship between time in Low-Level Blast Occupation and PHQ-9 ≥10 in age- and PTSD-adjusted models, with an estimated increase in the probability of clinical depression of roughly 0.24 (95% CrI ∼0.014-0.578) per one-standard-deviation increase in exposure via the mediator and a posterior probability P(IE>0) approaching 0.98. While PHQ-9 items overlap with sleep and affective symptoms, the threshold used here reflects clinically meaningful depression risk rather than diagnostic specificity, which is appropriate for an occupational readiness context.

A defining operational challenge is heterogeneity in that not all personnel with substantial blast histories develop depression, and group-level imaging effects can appear variable. Our moderation analyses provide a quantitative framework for this divergence by showing that the injury-outcome relationship depends on broader network context. At fixed high exposure, mediator-driven depression risk differed markedly across moderator configurations, producing zones of vulnerability and resilience. This suggests that “null” or modest average effects in prior studies may reflect averaging across individuals occupying different regions of the risk landscape rather than absence of injury. Given the dimensionality of the feature space and sample size, moderator findings should be interpreted as pattern-level evidence of heterogeneity rather than definitive effects of specific regions or measures.

These findings motivate a shift from static thresholds toward dynamic, individualized blast risk management. The mouse data challenge the sufficiency of single-event occupational limits by demonstrating that repetitive 3 psi exposures can yield chronic circuit-level injury when delivered in realistic training patterns. The human data highlight insula-centered diffusion/NODDI and Free-water features and network composites as candidate biomarkers that could be integrated with ongoing blast-dose monitoring to support early risk stratification. Finally, the heterogeneity framework suggests that prevention and mitigation strategies may be most effective when targeted to individuals with high-risk moderator configurations rather than applied uniformly.

Operationally, these results suggest a path toward precision blast medicine. First, exposure monitoring can move beyond peak-pressure thresholds by pairing cumulative impulse and training cadence with brain-based readouts that are mechanistically tied to mood risk. Second, insula-centered diffusion/NODDI features and network composites could serve as intermediate biomarkers for early identification of personnel whose brain network integrity is drifting into a high-risk state before overt clinical impairment. Third, the moderator-defined risk landscape implies that mitigation and care may be most effective when targeted such that individuals in vulnerability zones could receive prioritized sleep and mood interventions, dose-management counseling, and intensified monitoring, while resilient profiles may be safely maintained with standard surveillance. Finally, the mouse pathology highlights plausible biological mechanisms including neuroimmune activation, amyloid-positive vascular pathology and adjacent remodeling, and water-transport regulation that can be experimentally perturbed to test prevention strategies and to de-risk candidate therapeutics.

Several limitations warrant emphasis. Human analyses are cross-sectional and observational; longitudinal imaging and symptom tracking across careers and into retirement are required to establish temporal ordering and predictive validity. Sample size, while substantial for a deeply phenotyped SOF cohort, constrains stability of high-dimensional moderation screens and mandates replication. Time in Low-Level Blast Occupation remains a surrogate for individualized cumulative dose; future studies should integrate training records and blast-gauge data to derive individualized cumulative impulse or generalized blast exposure estimates and test whether the identified mediators and moderators retain predictive power.

In summary, by anchoring a sub-threshold 3 psi mouse model in monitored operational training exposure environments and integrating it with advanced neuroimaging and symptom phenotyping in U.S. Army SOF Soldiers, we demonstrate that repetitive LLB is associated with chronic disruption of an insula-centered cortico-striato-thalamic circuit across species. In SOF Soldiers, microstructural compromise of a right insula-basal ganglia network statistically mediates the association between cumulative occupational exposure and clinically significant depression risk, while network-level moderators help explain why comparable exposure histories can be accompanied by divergent clinical trajectories.

## Data Availability

All data produced in the present study are restricted from sharing without the approval and express written consent of the US Special Operations Command, the University of North Carolina at Chapel Hill and the Department of Veterans Affairs. Reasonable requests may be directed to the authors for initiation of those determinations.

## Author contributions

JSM, JPM: Supervision, funding acquisition; JSM, JPM, CM: Conceptualization, formal analysis, investigation, visualization, writing—all drafts, writing—review, editing, resources; RGT, TC, JWR, MO: Formal analysis, investigation; RGT, JSM: Validation; TLR and DL: Formal analysis, performed the DTI, NODDI, and assortativity analyses; TC, DRM, JWR, MO: Investigation; ERP, AC: Conceptualization, writing—revision, resources.

We thank C. Dirk Keene, Amanda Keen, Angela Wilson and Kim Howard for expert technical assistance. We also thank Daniel Corry, Brittany Heikke, and Kristen Schleich from the Matthew Gfeller Center at UNC for their assistance with compiling the raw human data analyzed in this manuscript. We are deeply grateful to the military Servicemembers, Veterans, civilian participants, and their families whose participation made this work possible. Figure 1a was created in BioRender. Meabon, J. (2026) https://BioRender.com/z19q670

## Funding

This work was supported by the Veterans Affairs Office of Biomedical Laboratory Research and Development (J.S.M., I01BX004896), the University of Washington Friends of Alzheimers Research (E.R.P.), the VA Northwest Mental Illness Research, Education and Clinical Center (J.S.M., E.R.P.) and a Congressionally Directed Medical Research TBI Psychological Health Research Program award (J.S.M., J.P.M.; HT94252310755). The funding agencies had no role in study design, data collection, data analysis, data interpretation or writing of the manuscript. The contents are solely the responsibility of the authors and do not necessarily represent the views of the US Department of Veterans Affairs, the US Department of Defense, the United States Government or any affiliated agencies. The human data used in this project were collected as part of the Assessing & Tracking Tactical (ATTAC) Forces Initiative originally supported by the US Army Medical Research and Development Command under Contract No. W81XWH-20-C-0022 and continued under an effort sponsored by the Government under Other Transactions Number W81XWH-15-9-0001. The views and conclusions contained herein are those of the authors and should not be interpreted as necessarily representing the official policies or endorsements, either expressed or implied, of the U.S. Government. The Public Affairs Office at the United States Special Operations Command and the United States Army Special Operations Command approved this submission for public review and release.

## Competing Interests

The authors declare no conflict of interest with the present study.

## Supplemental Materials

**Supplemental Figure 1.**
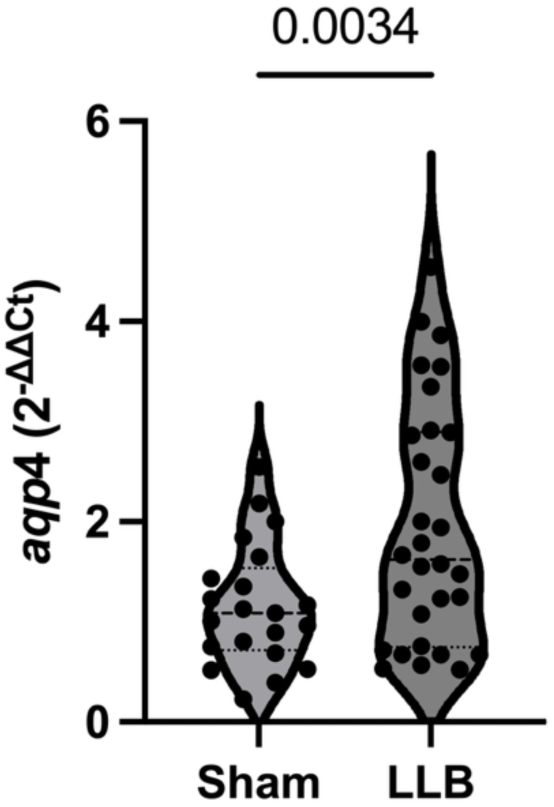
Repeated low-level blast increases *apq*4 levels in frontal cortex in mice. Mean aquaporin-4 mRNA expression in individual study mice (dots) aged 6-months after LLB/sham exposures. Each dot is the mean of triplicate technical replicates from N=21 sham, N=30 LLB mice. 2-tailed Welch’s *t*-test.

**Supplemental Figure 2.**
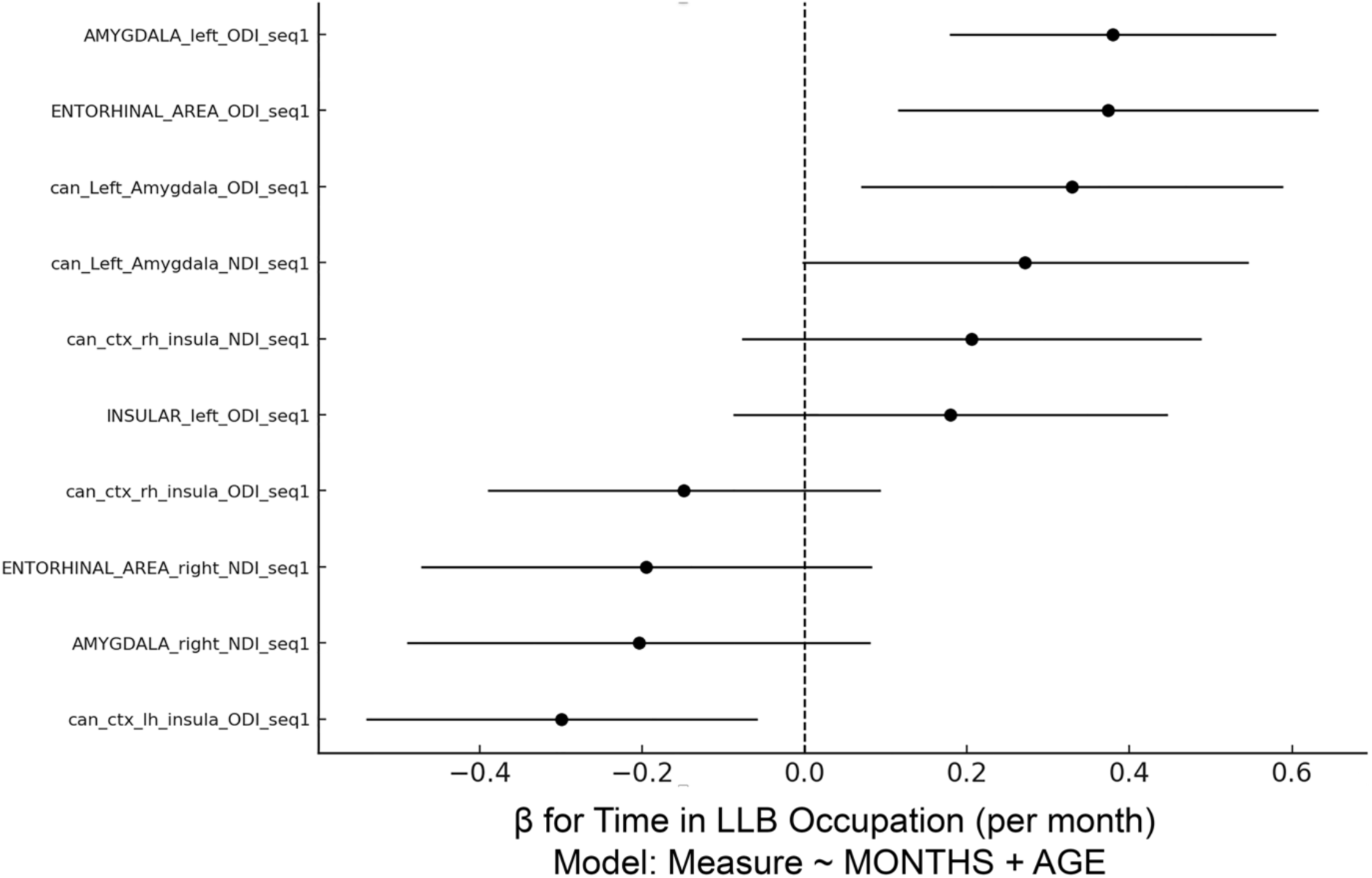
ODI/NDI vs. Time in LLB Occupation (x-axis). Among the top ten by posterior evidence, four show ≥95% probability of a positive months effect and one shows ≥95% probability of a negative effect; at the ≥90% threshold, positives=6, negatives=3. Directionality implies that with longer Time in LLB Occupation (as a proxy for cumulative low-level blast exposure), positively changing ODI/NDI likely increase, while negatively changing measures likely decrease.

**Supplemental Table 1.** Demographic and exposure characteristics.

| Variable | Trainees (n=31) | New SOF (n=55) | Experienced SOF (n=51) | Retired SOF (n=10) |
| --- | --- | --- | --- | --- |
| **Time in LLB Occupation (months)** | — | 0.0 ± 0.0 [0.0–0.0]; reported 67.3% (37/55) | 56.1 ± 27.0 [9.0–127.0]; reported 54.9% (28/51) | —; reported 0.0% (0/10) |
| **Age (years)** | — | 31.6 ± 3.5 [24.0–38.0]; reported 67.3% (37/55) | 36.9 ± 5.7 [22.0–54.0]; reported 78.4% (40/51) | 46.0 ± — [46.0–46.0]; reported 10.0% (1/10) |
| **History of concussion (self-report)** | reported 100.0% (31/31) | reported 78.2% (43/55) | reported 54.9% (28/51) | reported 10.0% (1/10) |
| - Yes | 77.4% (24/31) | 65.5% (36/55) | 49.0% (25/51) | 10.0% (1/10) |
| - Unsure/Don't know | 12.9% (4/31) | 7.3% (4/55) | 3.9% (2/51) | 0.0% (0/10) |
| - No | 9.7% (3/31) | 5.5% (3/55) | 2.0% (1/51) | 0.0% (0/10) |
| **Number of concussions (self-report)** | —; reported 0.0% (0/31) | 3 ± 2.7 [1–10]; reported 30.9% (17/55) | 4 ± 2.9 [1–10]; reported 54.9% (28/51) | 3 ± — [3–3]; reported 10.0% (1/10) |
| **Age at enlistment (years)** | —; reported 0.0% (0/31) | 21.1 ± 2.7 [18.0–27.0]; reported 30.9% (17/55) | 19.9 ± 2.3 [17.0–24.0]; reported 35.3% (18/51) | —; reported 0.0% (0/10) |
| **Height** | 5' 11" ± 2.6" [5' 6"–6' 4"]; reported 100.0% (31/31) | 5' 11" ± 2.6" [5' 6"–6' 5"]; reported 78.2% (43/55) | 5' 11" ± 2.2" [5' 7"–6' 4"]; reported 54.9% (28/51) | 6' 1" ± — [6' 1"–6' 1"]; reported 10.0% (1/10) |
| **Weight (lb)** | 187.5 ± 15.1 [159.0–220.0]; reported 100.0% (31/31) | 186.7 ± 19.8 [135.0–235.0]; reported 78.2% (43/55) | 194.3 ± 16.5 [170.0–225.0]; reported 54.9% (28/51) | 245.0 ± — [245.0–245.0]; reported 10.0% (1/10) |
| **Hispanic** | reported 100.0% (31/31) | reported 78.2% (43/55) | reported 54.9% (28/51) | reported 10.0% (1/10) |
| - Not Hispanic/Latino | 83.9% (26/31) | 60.0% (33/55) | 47.1% (24/51) | 10.0% (1/10) |
| - Hispanic/Latino | 9.7% (3/31) | 9.1% (5/55) | 2.0% (1/51) | 0.0% (0/10) |
| - Unknown | 6.5% (2/31) | 3.6% (2/55) | 3.9% (2/51) | 0.0% (0/10) |
| - Prefer not to answer | 0.0% (0/31) | 5.5% (3/55) | 2.0% (1/51) | 0.0% (0/10) |
| **Race** | reported 100.0% (31/31) | reported 78.2% (43/55) | reported 54.9% (28/51) | reported 10.0% (1/10) |
| - White | 93.5% (29/31) | 65.5% (36/55) | 51.0% (26/51) | 10.0% (1/10) |
| - Other | 3.2% (1/31) | 3.6% (2/55) | 2.0% (1/51) | 0.0% (0/10) |
| - Native Hawaiian/Pacific Islander | 3.2% (1/31) | 1.8% (1/55) | 0.0% (0/51) | 0.0% (0/10) |
| - American Indian/Alaska Native | 0.0% (0/31) | 3.6% (2/55) | 0.0% (0/51) | 0.0% (0/10) |
| - Prefer not to answer | 0.0% (0/31) | 1.8% (1/55) | 0.0% (0/51) | 0.0% (0/10) |
| - More than one race | 0.0% (0/31) | 1.8% (1/55) | 0.0% (0/51) | 0.0% (0/10) |
| - African American/Black | 0.0% (0/31) | 0.0% (0/55) | 2.0% (1/51) | 0.0% (0/10) |
| **Declared Marital Status** | reported 100.0% (31/31) | reported 78.2% (43/55) | reported 54.9% (28/51) | reported 10.0% (1/10) |
| - Married | 77.4% (24/31) | 60.0% (33/55) | 47.1% (24/51) | 10.0% (1/10) |
| - Single (never married) | 12.9% (4/31) | 10.9% (6/55) | 2.0% (1/51) | 0.0% (0/10) |
| - Separated/Divorced | 6.5% (2/31) | 5.5% (3/55) | 5.9% (3/51) | 0.0% (0/10) |
| - Living with significant other | 3.2% (1/31) | 1.8% (1/55) | 0.0% (0/51) | 0.0% (0/10) |
| **Education** | reported 100.0% (31/31) | reported 78.2% (43/55) | reported 54.9% (28/51) | reported 10.0% (1/10) |
| - Some college | 32.3% (10/31) | 29.1% (16/55) | 19.6% (10/51) | 10.0% (1/10) |
| - Bachelor's degree | 35.5% (11/31) | 25.5% (14/55) | 7.8% (4/51) | 0.0% (0/10) |
| - Post-Graduate school or degree | 16.1% (5/31) | 10.9% (6/55) | 5.9% (3/51) | 0.0% (0/10) |
| - Associates degree | 9.7% (3/31) | 9.1% (5/55) | 7.8% (4/51) | 0.0% (0/10) |
| - High School Graduate - high school diploma or equivalent | 6.5% (2/31) | 3.6% (2/55) | 13.7% (7/51) | 0.0% (0/10) |
| **Rank** | reported 100.0% (31/31) | reported 78.2% (43/55) | reported 54.9% (28/51) | reported 10.0% (1/10) |
| - E7 | 51.6% (16/31) | 34.5% (19/55) | 9.8% (5/51) | 0.0% (0/10) |
| - E8 | 3.2% (1/31) | 16.4% (9/55) | 37.3% (19/51) | 0.0% (0/10) |
| - E6 | 22.6% (7/31) | 20.0% (11/55) | 0.0% (0/51) | 0.0% (0/10) |
| - O4 | 16.1% (5/31) | 5.5% (3/55) | 0.0% (0/51) | 0.0% (0/10) |
| - E9 | 0.0% (0/31) | 0.0% (0/55) | 7.8% (4/51) | 10.0% (1/10) |
| - O3 | 6.5% (2/31) | 1.8% (1/55) | 0.0% (0/51) | 0.0% (0/10) |
| **Helmet use** | reported 96.8% (30/31) | reported 78.2% (43/55) | reported 54.9% (28/51) | reported 10.0% (1/10) |
| - Always | 90.3% (28/31) | 72.7% (40/55) | 52.9% (27/51) | 10.0% (1/10) |
| - Sometimes | 6.5% (2/31) | 5.5% (3/55) | 2.0% (1/51) | 0.0% (0/10) |
| **Body armor** | reported 96.8% (30/31) | reported 78.2% (43/55) | reported 54.9% (28/51) | reported 10.0% (1/10) |
| - Always | 96.8% (30/31) | 76.4% (42/55) | 52.9% (27/51) | 10.0% (1/10) |
| - Sometimes | 0.0% (0/31) | 1.8% (1/55) | 2.0% (1/51) | 0.0% (0/10) |
| **Eye protection** | reported 96.8% (30/31) | reported 78.2% (43/55) | reported 54.9% (28/51) | reported 10.0% (1/10) |
| - Always | 74.2% (23/31) | 74.5% (41/55) | 51.0% (26/51) | 10.0% (1/10) |
| - Sometimes | 19.4% (6/31) | 3.6% (2/55) | 3.9% (2/51) | 0.0% (0/10) |
| - Never | 3.2% (1/31) | 0.0% (0/55) | 0.0% (0/51) | 0.0% (0/10) |
| **Passive hearing protection** | reported 96.8% (30/31) | reported 78.2% (43/55) | reported 52.9% (27/51) | reported 10.0% (1/10) |
| - Never | 51.6% (16/31) | 30.9% (17/55) | 33.3% (17/51) | 10.0% (1/10) |
| - Sometimes | 25.8% (8/31) | 36.4% (20/55) | 9.8% (5/51) | 0.0% (0/10) |
| - Always | 19.4% (6/31) | 9.1% (5/55) | 7.8% (4/51) | 0.0% (0/10) |
| - Does not apply | 0.0% (0/31) | 1.8% (1/55) | 2.0% (1/51) | 0.0% (0/10) |
| **Active hearing protection** | reported 96.8% (30/31) | reported 78.2% (43/55) | reported 54.9% (28/51) | reported 10.0% (1/10) |
| - Always | 77.4% (24/31) | 74.5% (41/55) | 52.9% (27/51) | 10.0% (1/10) |
| - Sometimes | 19.4% (6/31) | 3.6% (2/55) | 2.0% (1/51) | 0.0% (0/10) |
| **Tobacco use** | reported 100.0% (31/31) | reported 78.2% (43/55) | reported 54.9% (28/51) | reported 10.0% (1/10) |
| - No | 87.1% (27/31) | 58.2% (32/55) | 35.3% (18/51) | 10.0% (1/10) |
| - Yes | 12.9% (4/31) | 18.2% (10/55) | 19.6% (10/51) | 0.0% (0/10) |
| - Prefer not to answer | 0.0% (0/31) | 1.8% (1/55) | 0.0% (0/51) | 0.0% (0/10) |
| **Alcohol use days per month** | 2.7 ± 2.5 [0.0–10.0]; reported 100.0% (31/31) | 4.7 ± 4.1 [0.0–21.0]; reported 74.5% (41/55) | 5.4 ± 4.5 [0.0–15.0]; reported 54.9% (28/51) | 6.0 ± — [6.0–6.0]; reported 10.0% (1/10) |

**Supplemental Table 2.** Bayesian posterior evidence for ODI/NDI vs Time in LLB Occupation relationships.

| Region | measure | $\beta_{\text{time}}$ | CI95_low | CI95_hi | $p_{\text{time}}$ | Post prob $\beta < 0$ | Post prob $\beta > 0$ | Direction | N |
| --- | --- | --- | --- | --- | --- | --- | --- | --- | --- |
| Amygdala_left | ODI | 0.3794 | 0.1787 | 0.5802 | 0.000351 | 1.000 | 0.000 | positive | 67 |
| Entorhinal_left | ODI | 0.3737 | 0.1151 | 0.6324 | 0.00531 | 0.997 | 0.003 | positive | 67 |
| Amygdala_left | ODI | 0.3292 | 0.0695 | 0.5889 | 0.0138 | 0.993 | 0.007 | positive | 67 |
| Insula_left | ODI | -0.2993 | -0.5403 | -0.0583 | 0.0157 | 0.008 | 0.992 | negative | 67 |
| Amygdala_left | NDI | 0.2716 | -0.0031 | 0.5463 | 0.0525 | 0.974 | 0.026 | positive | 67 |
| Insula_right | NDI | 0.2058 | -0.0773 | 0.4888 | 0.151 | 0.924 | 0.076 | Likely positive | 67 |
| Amygdala_right | NDI | -0.2044 | -0.4896 | 0.0807 | 0.157 | 0.078 | 0.922 | Likely negative | 67 |
| Entorhinal_right | NDI | -0.1948 | -0.4728 | 0.0832 | 0.166 | 0.083 | 0.917 | Likely negative | 67 |
| Insula_left | ODI | 0.1793 | -0.0881 | 0.4468 | 0.185 | 0.907 | 0.093 | Likely positive | 67 |
| Insula_right | ODI | -0.1485 | -0.3906 | 0.0936 | 0.225 | 0.112 | 0.888 | Likely negative | 67 |

## Supplemental Information 1. Code Scripts

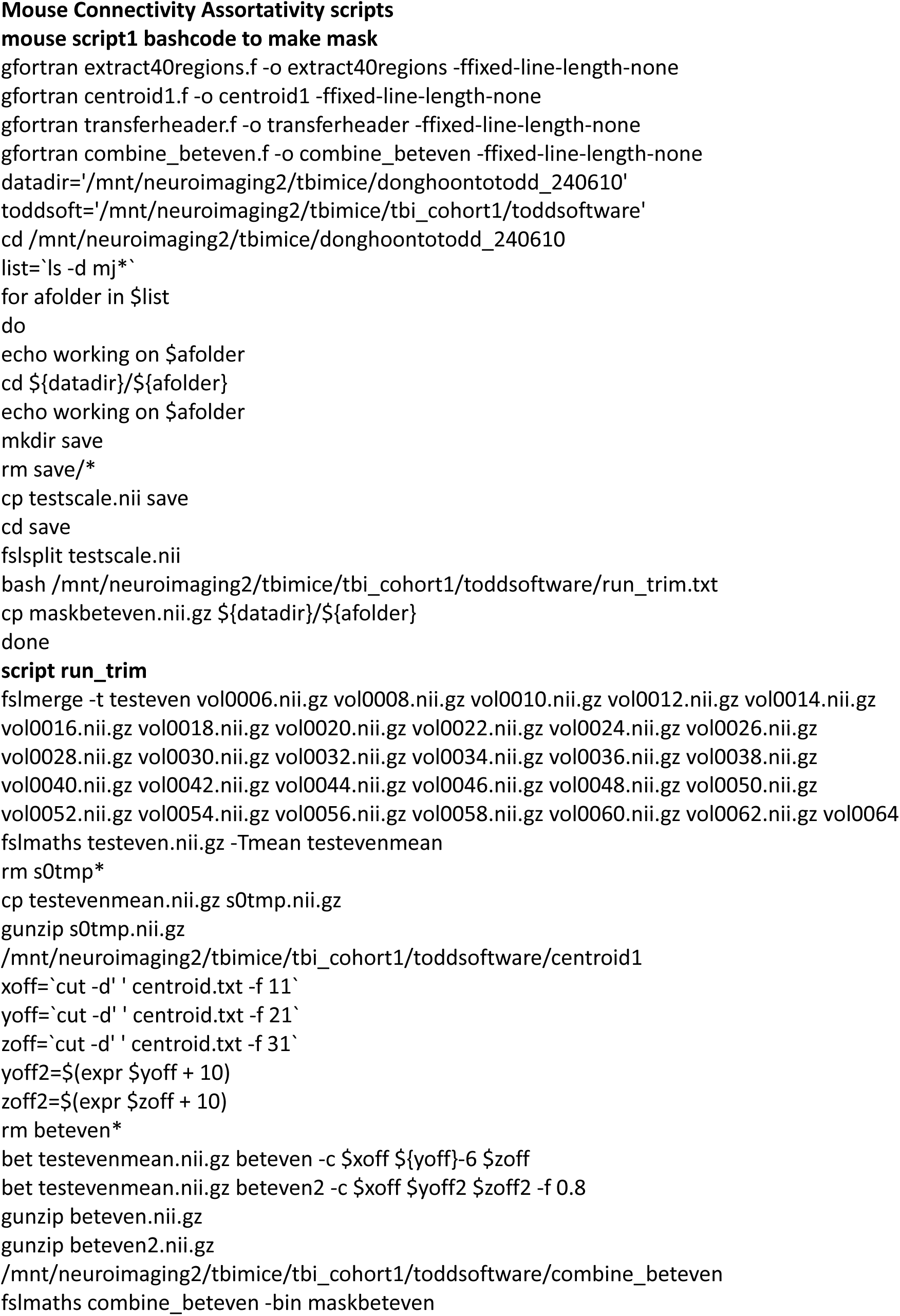
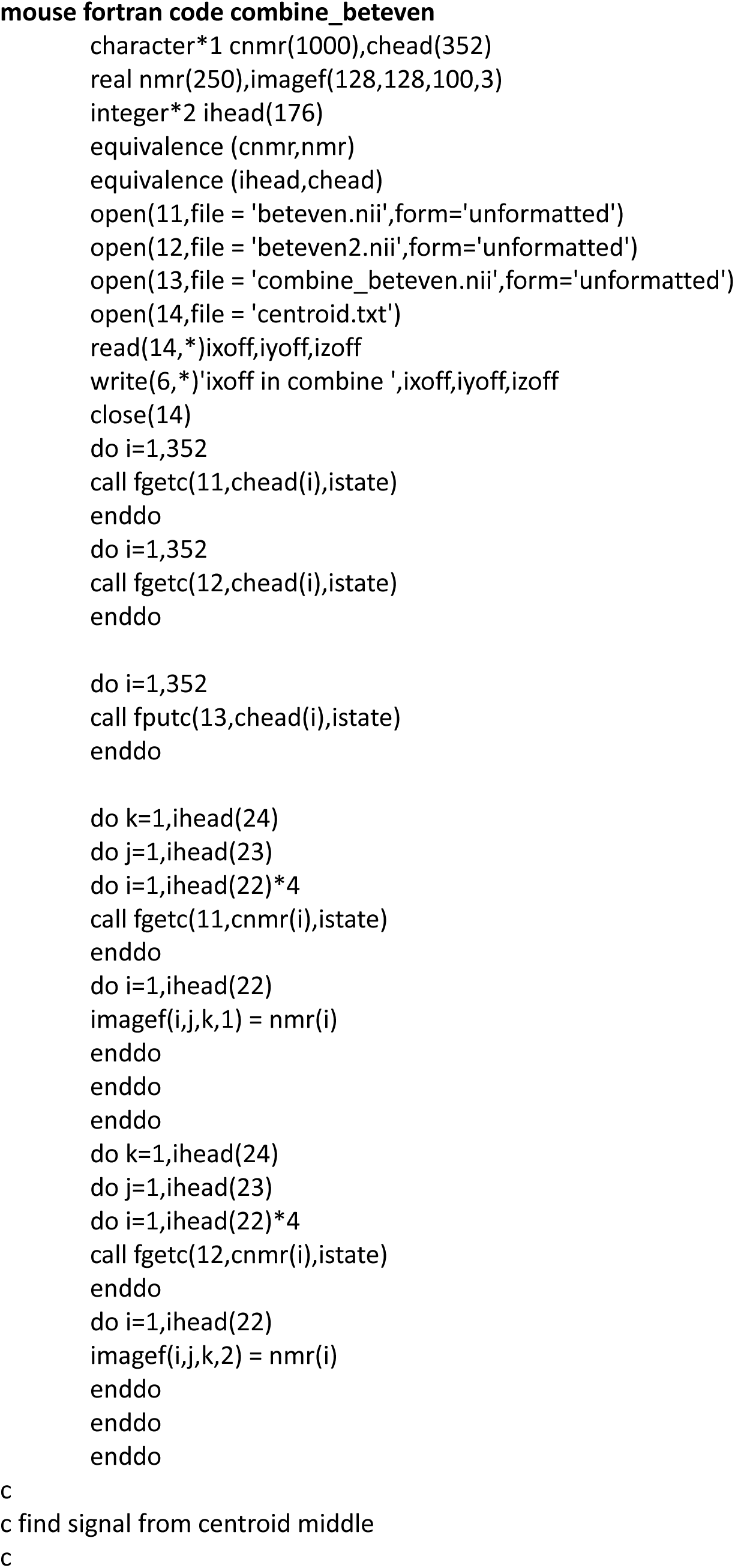
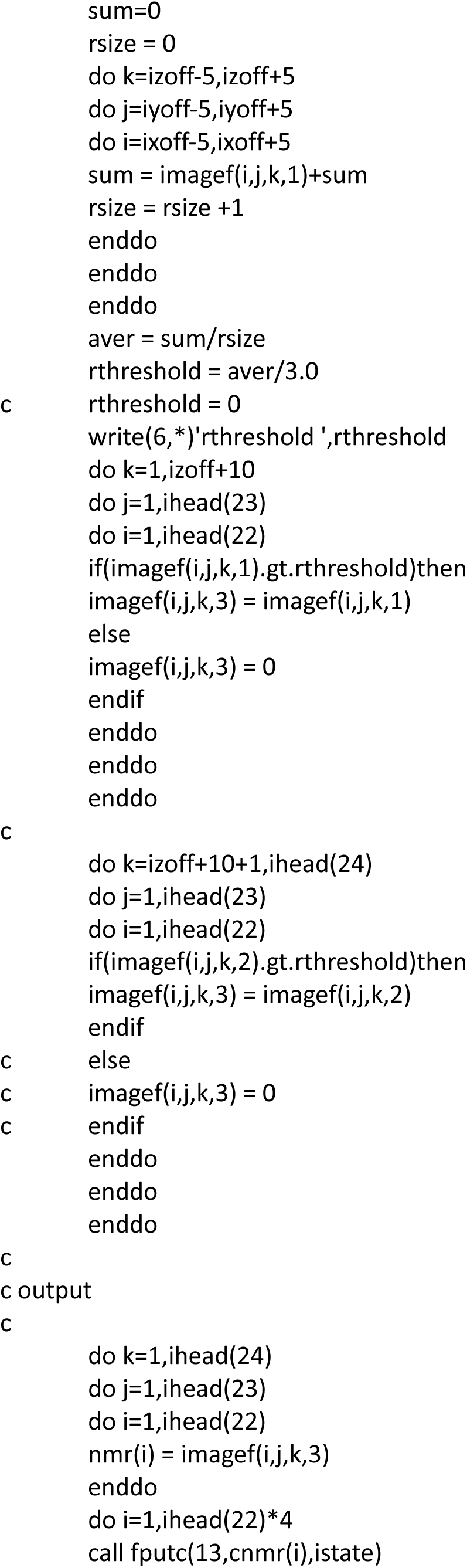
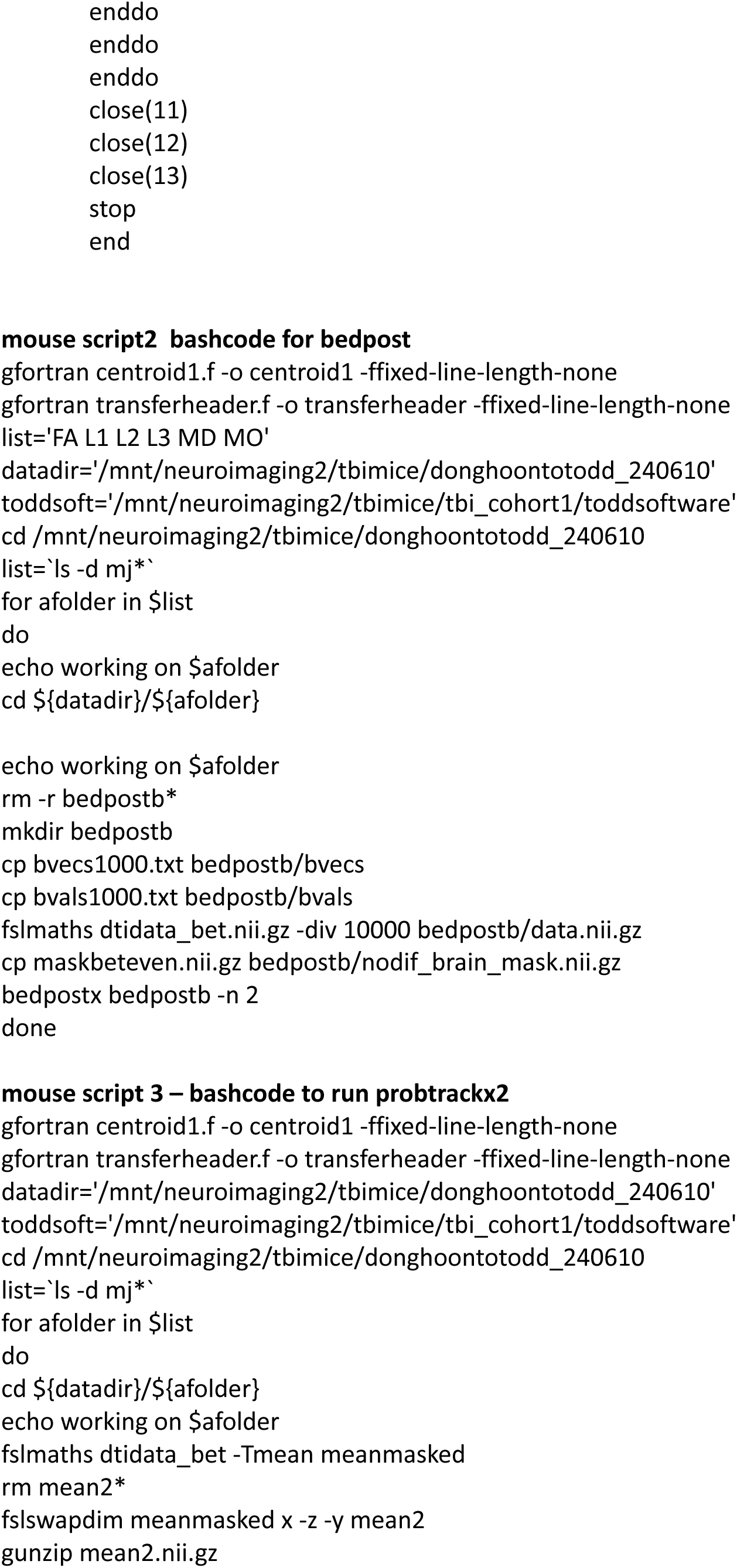
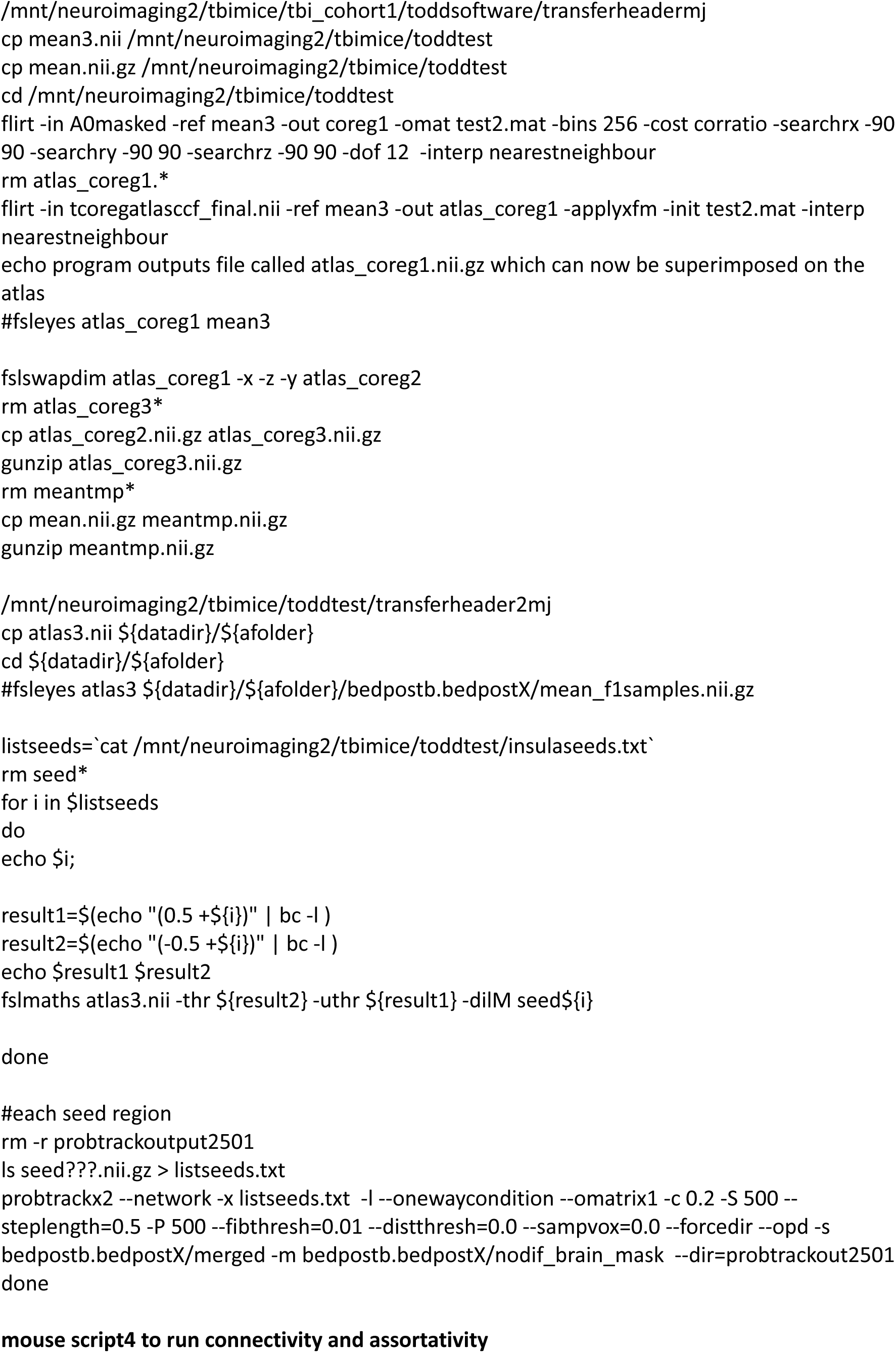
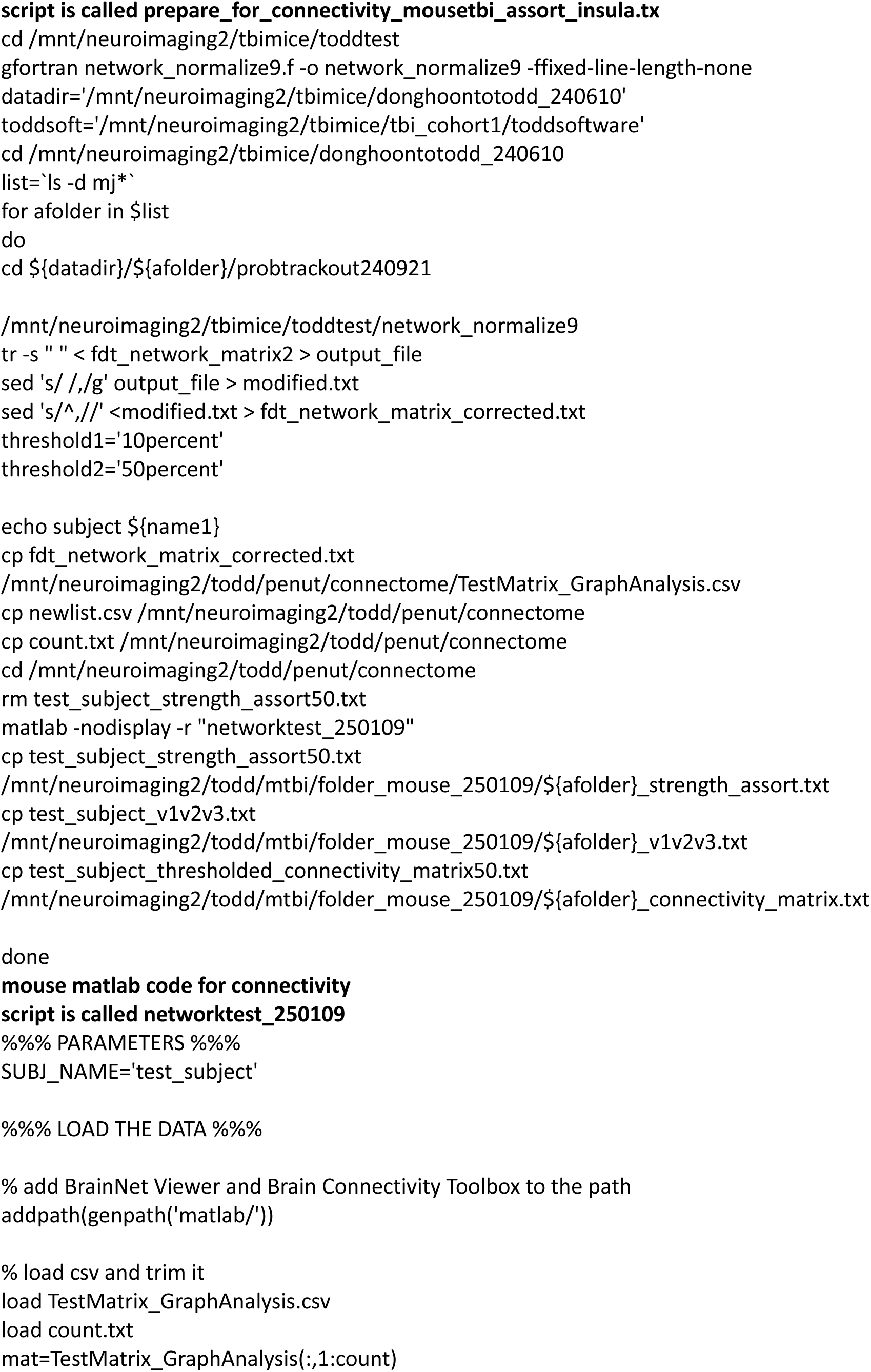
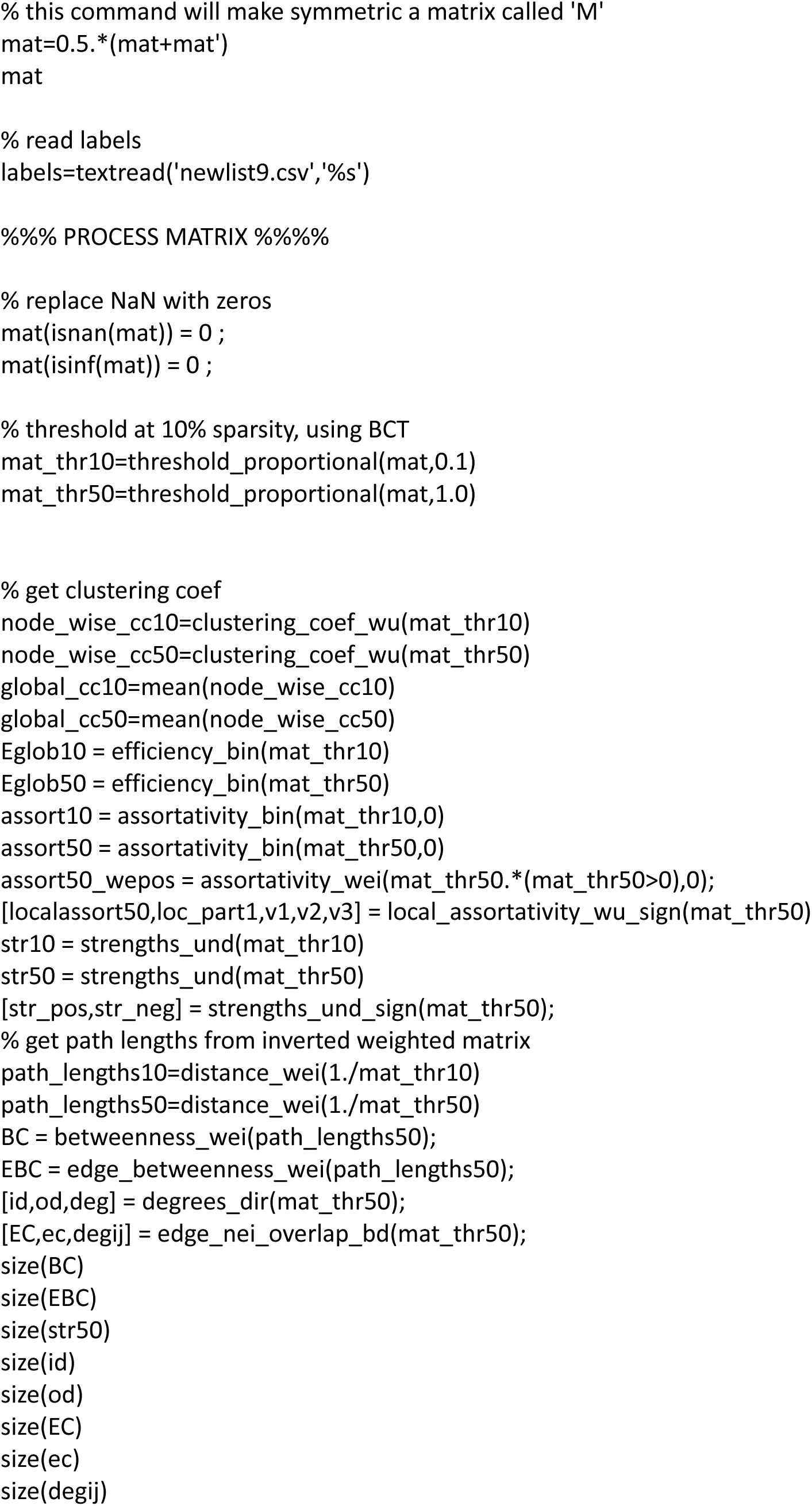
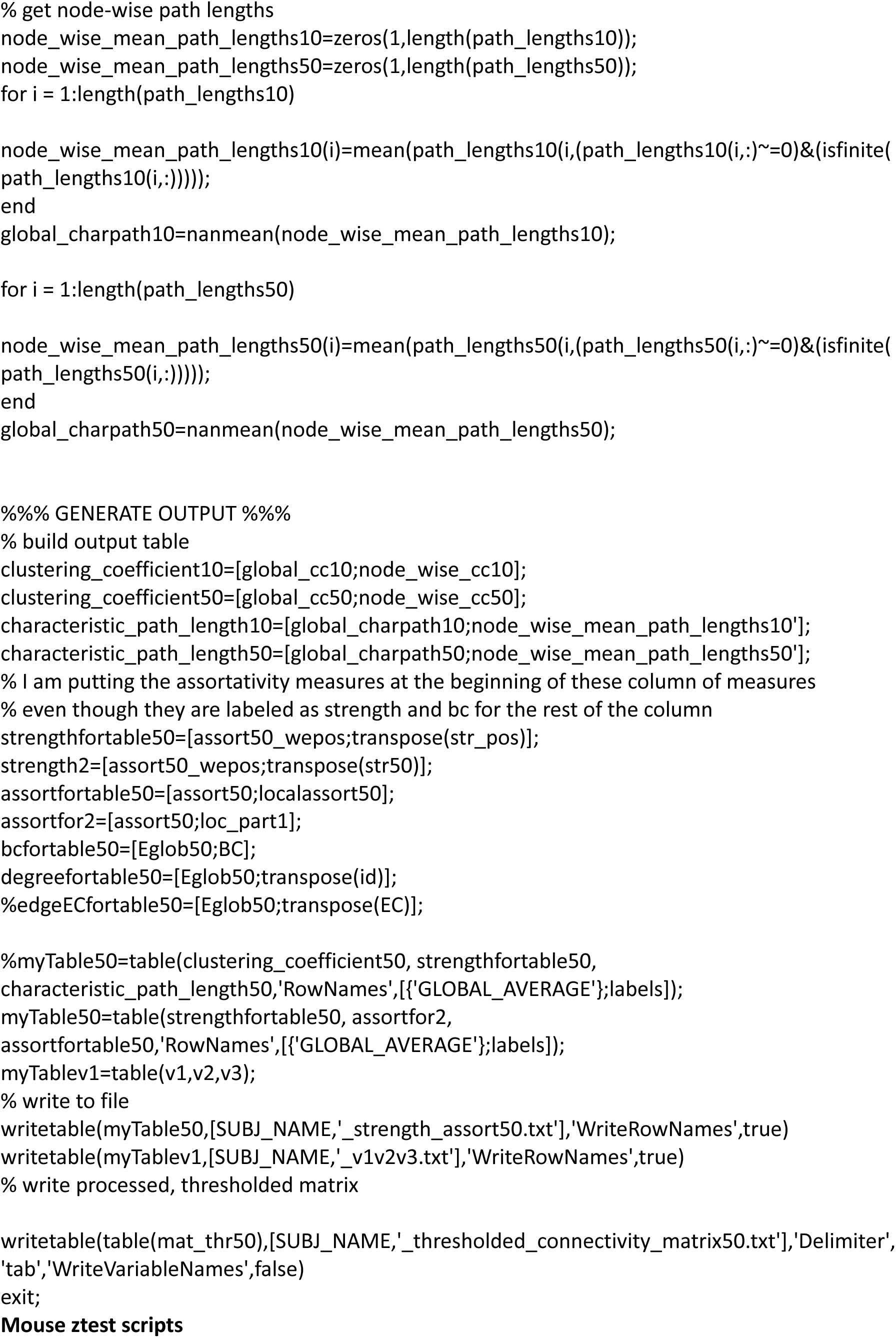
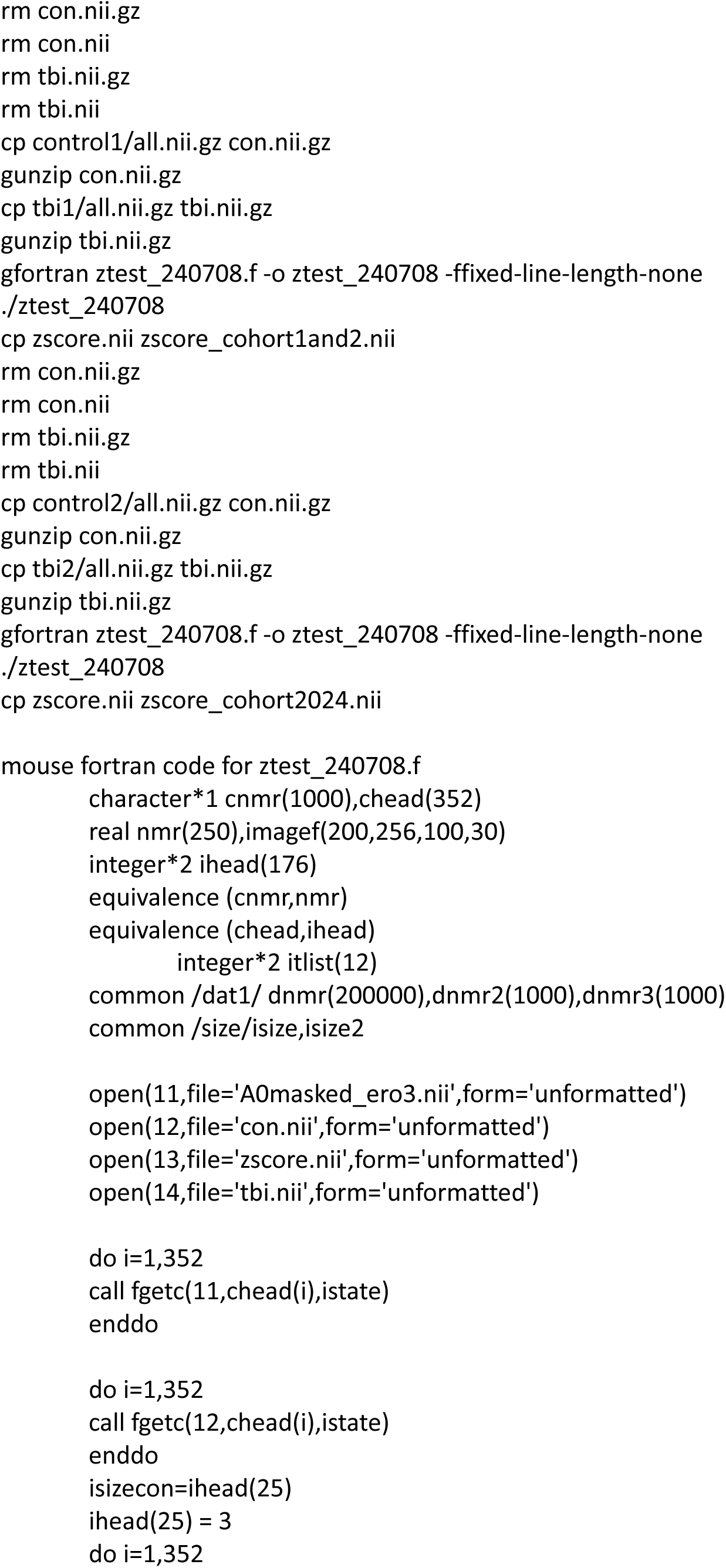
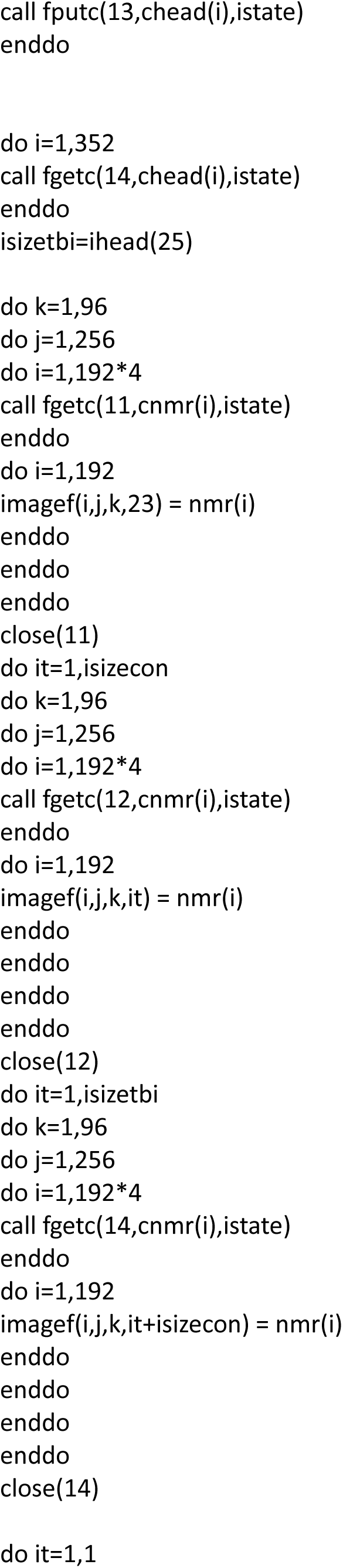
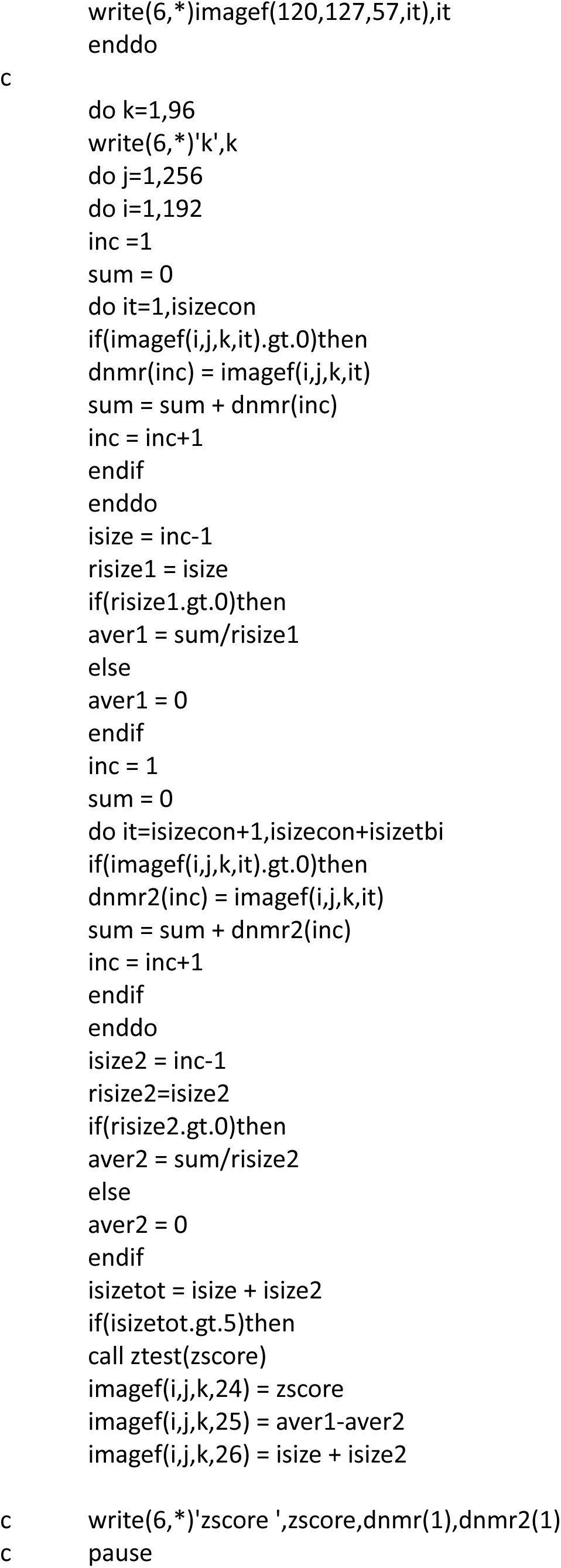
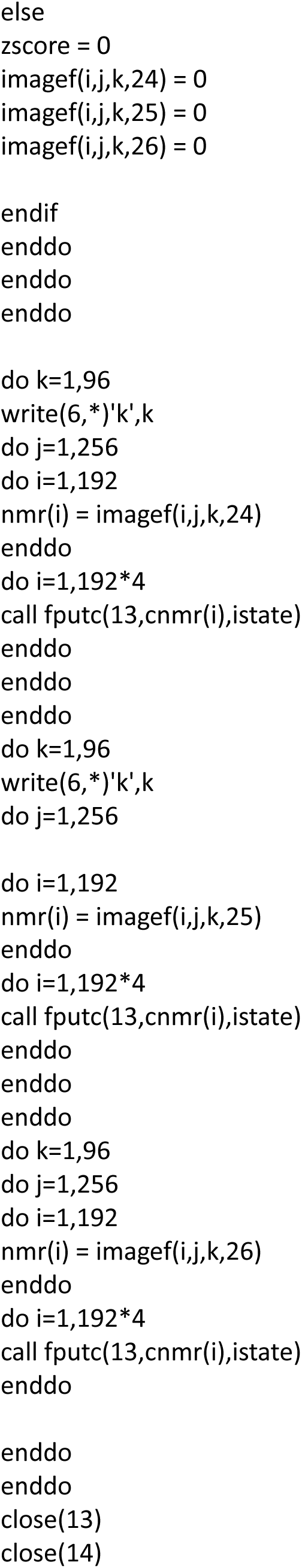
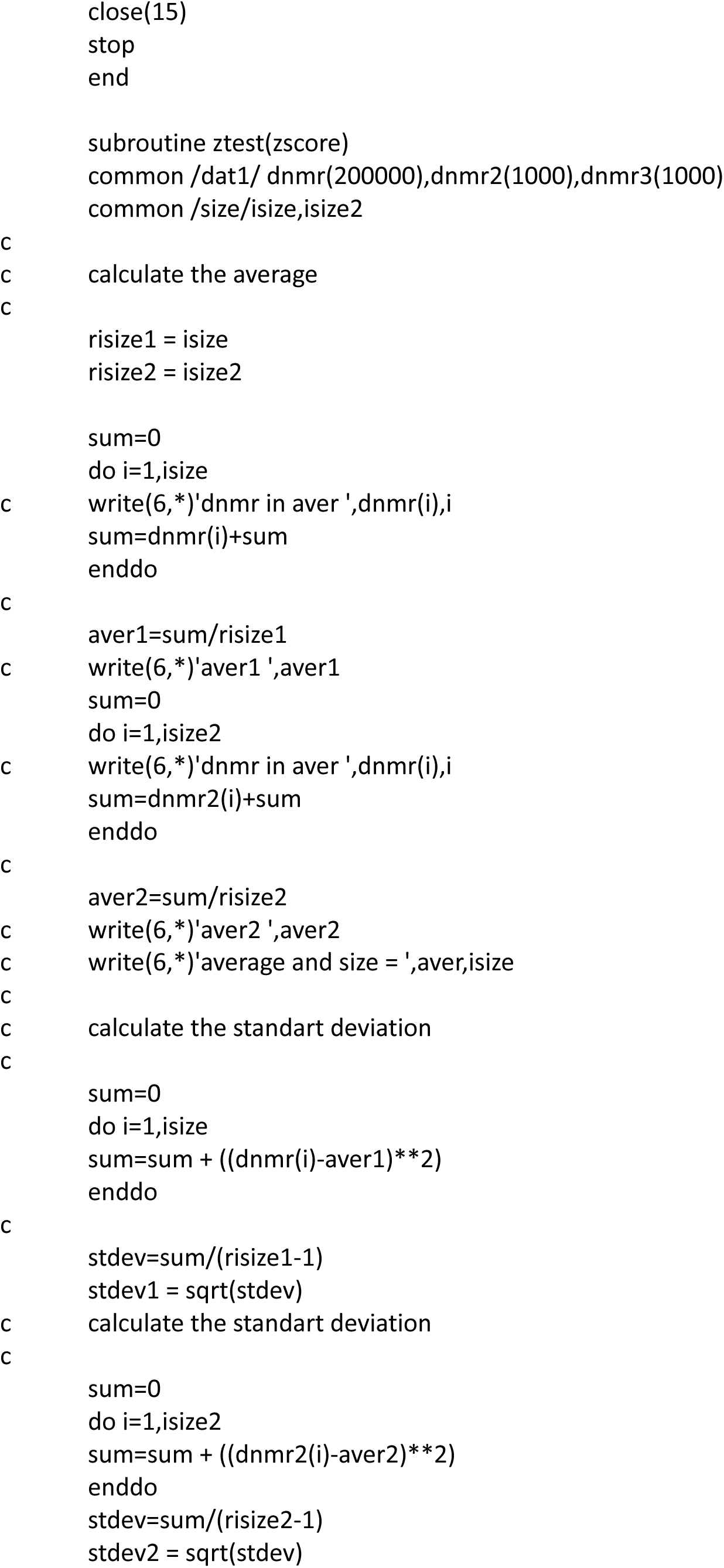
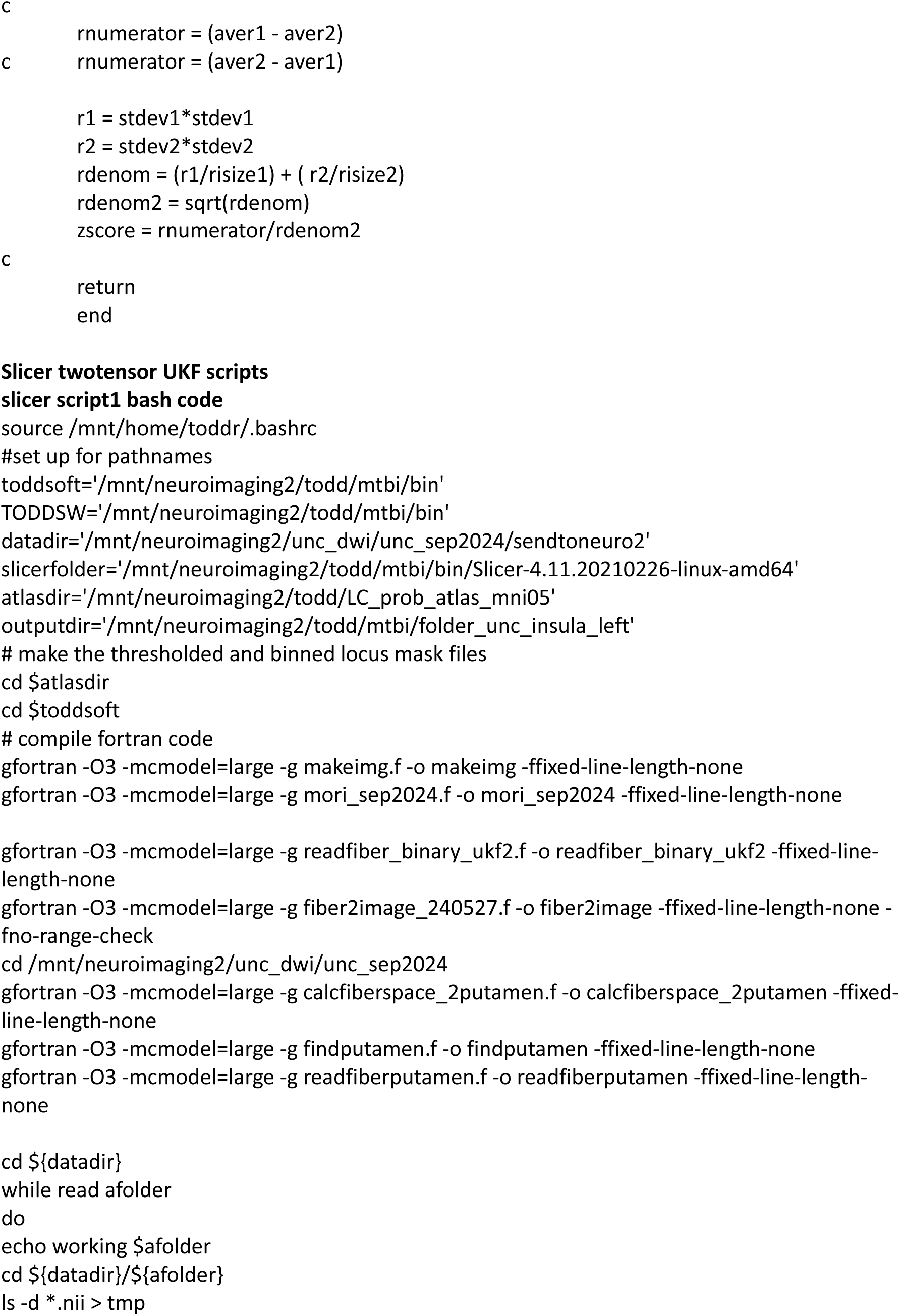
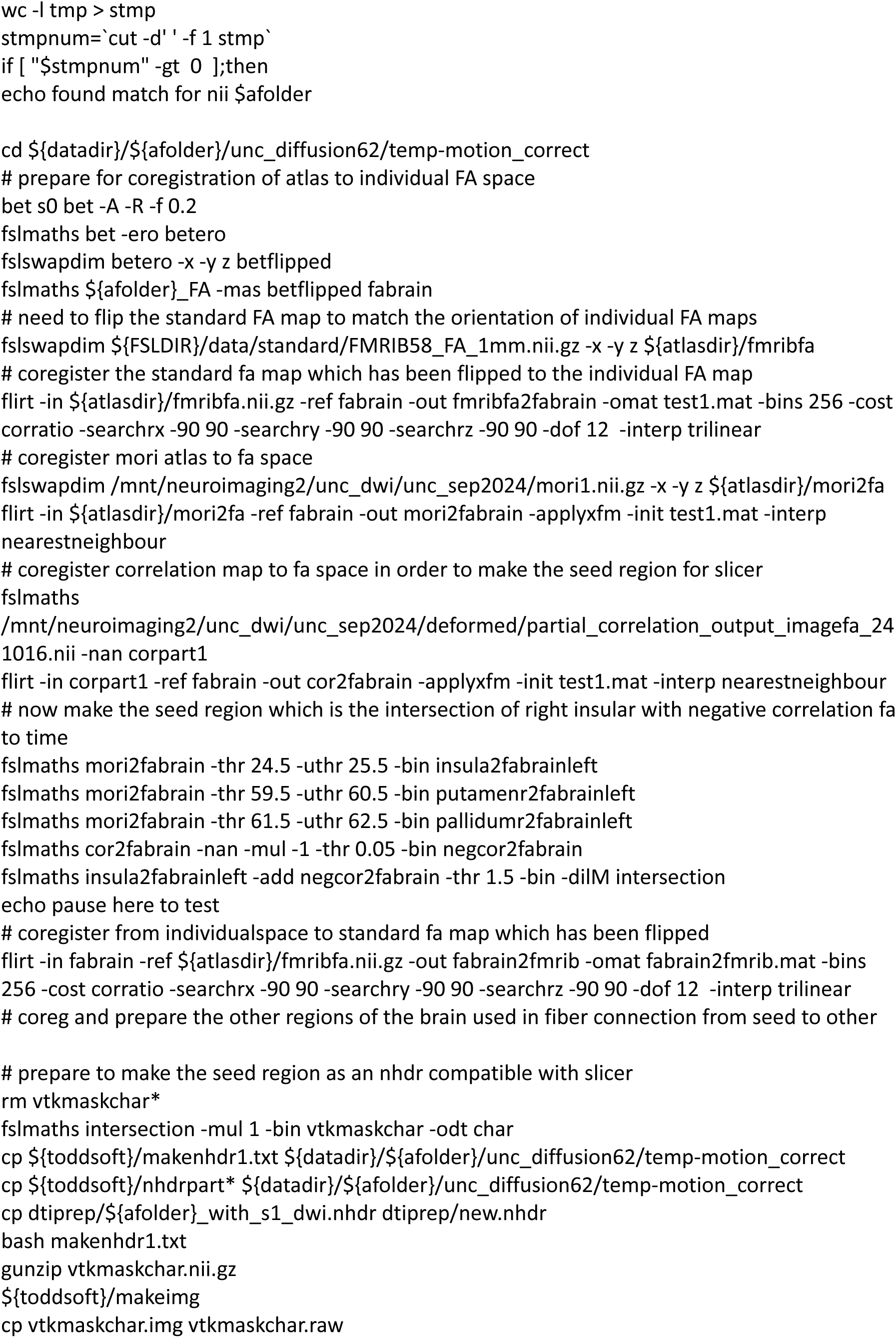
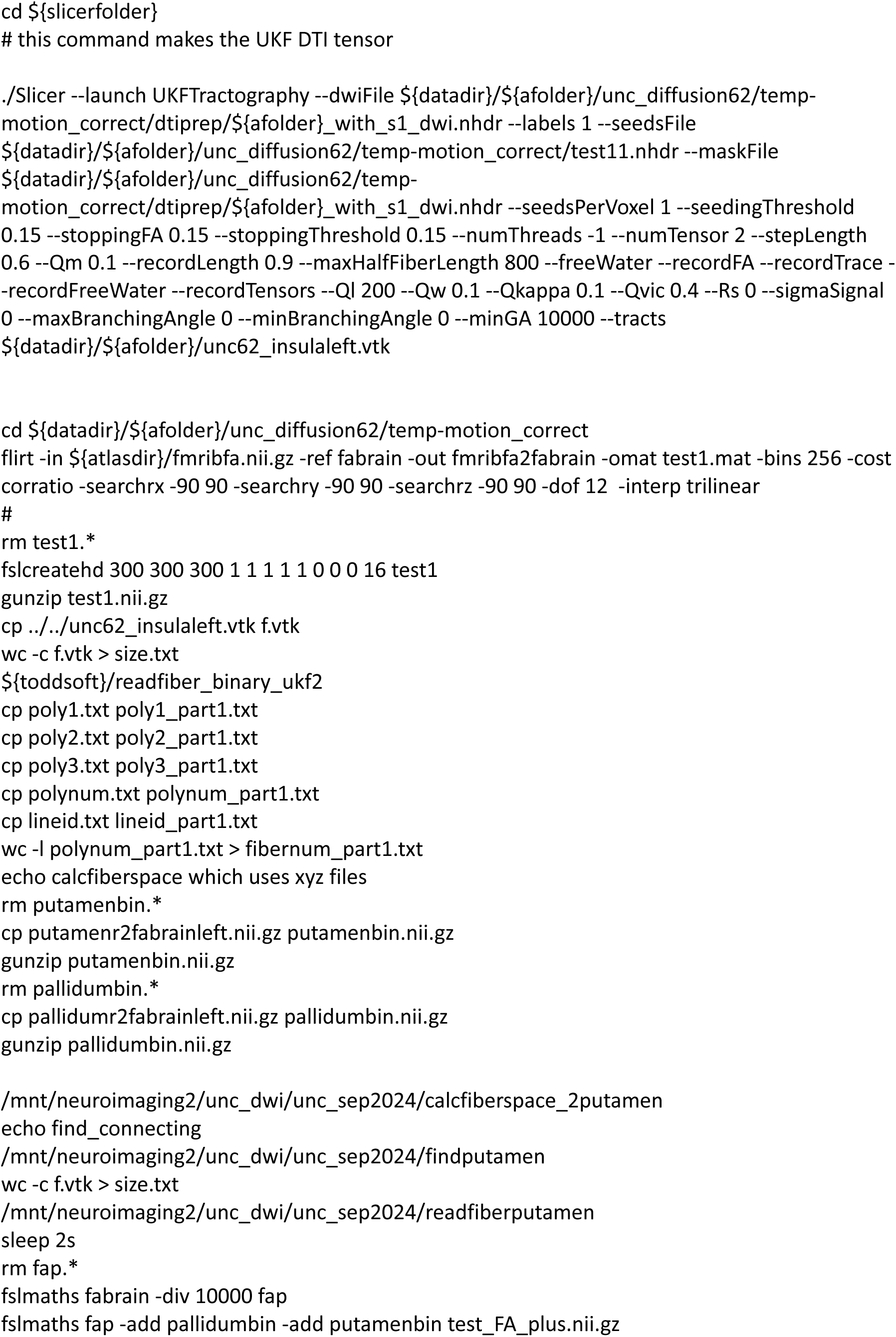
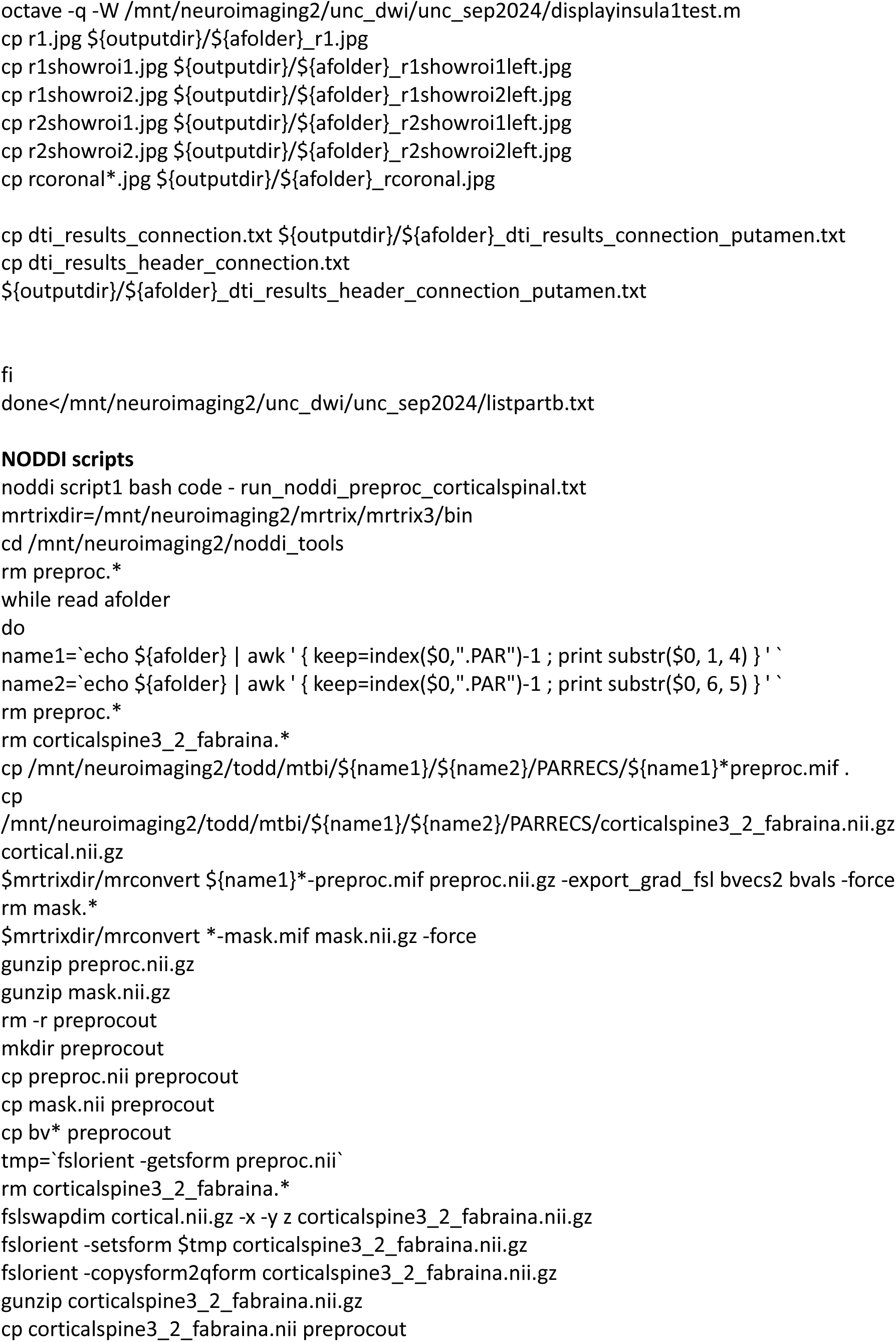
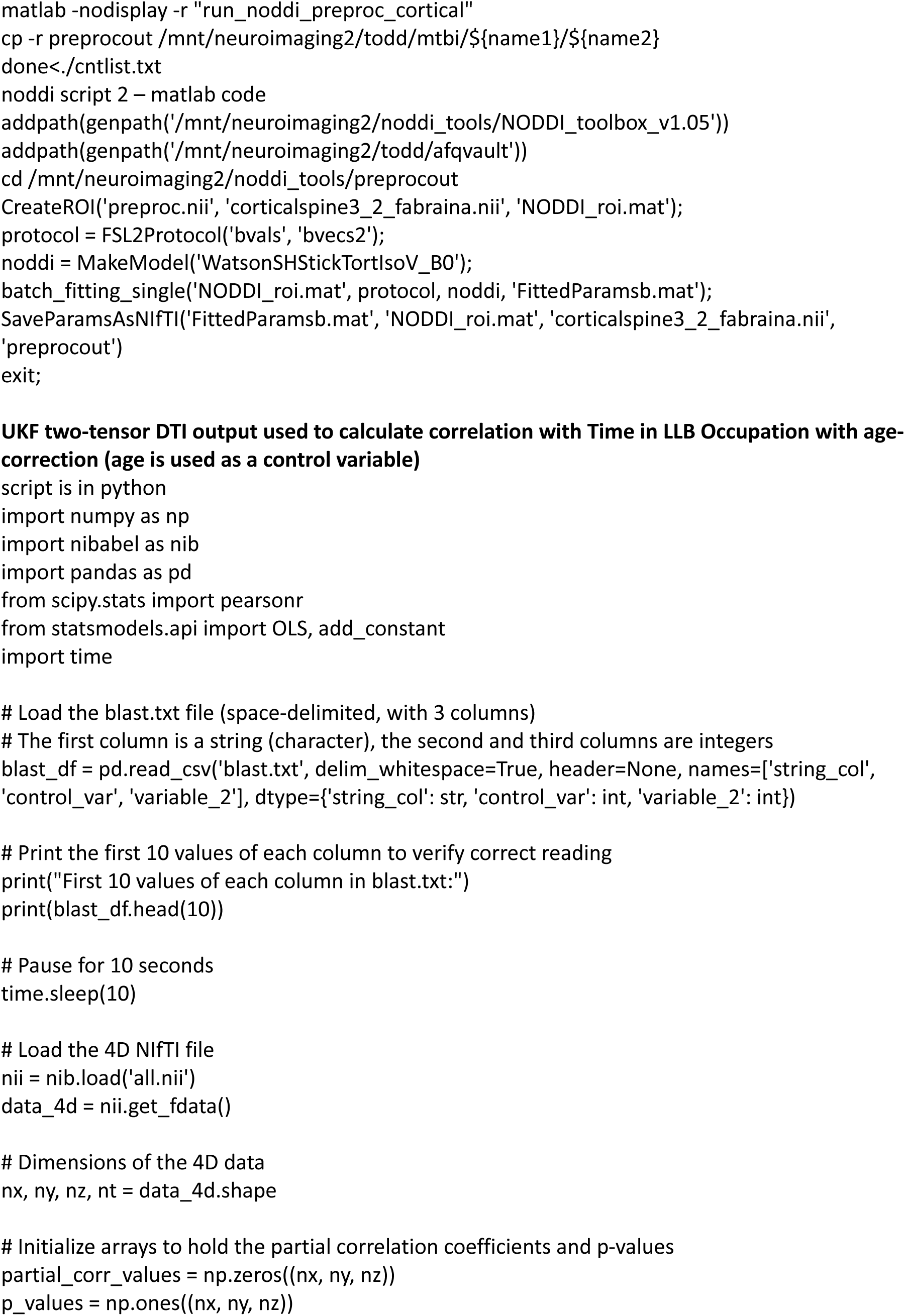
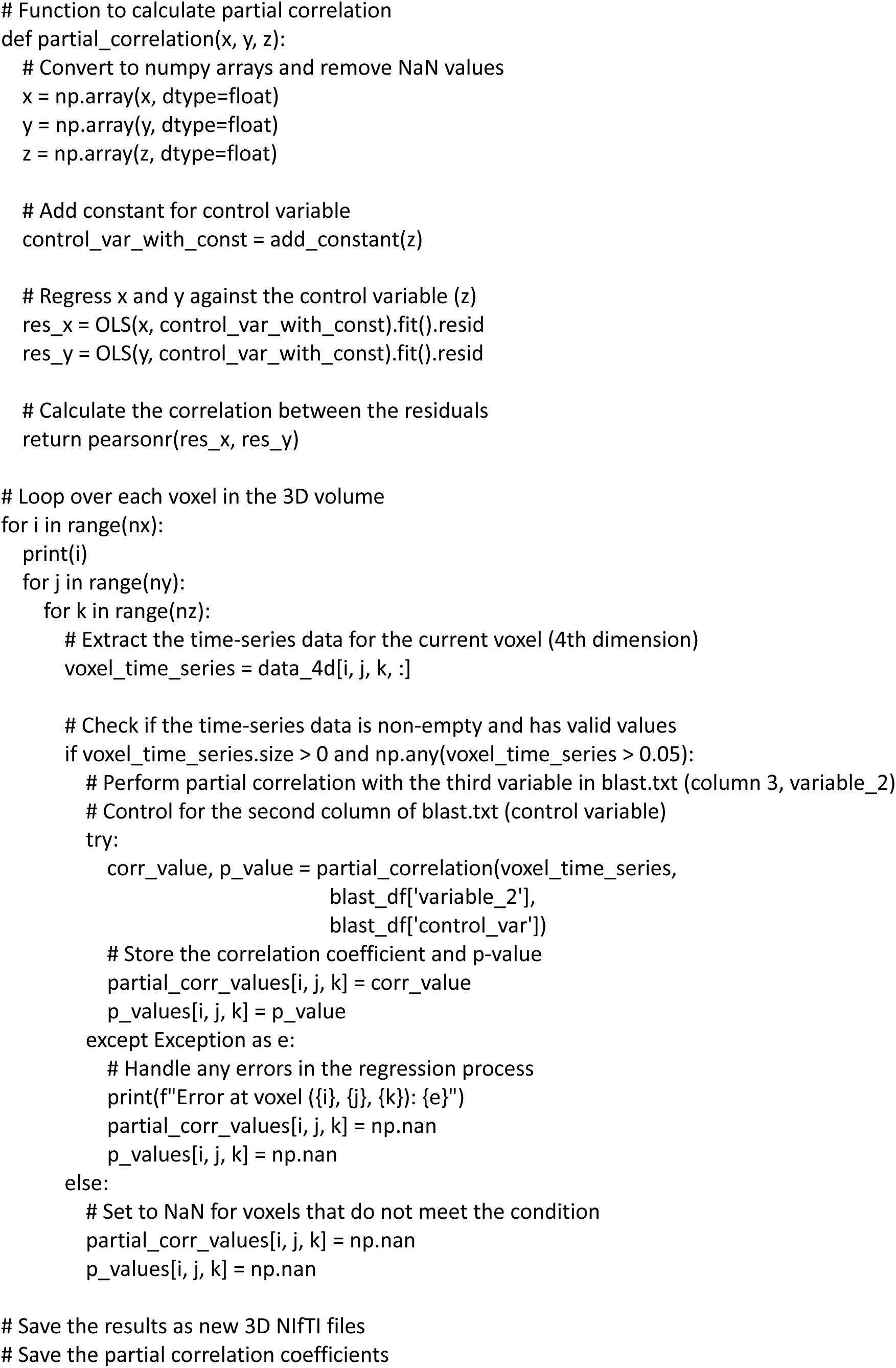
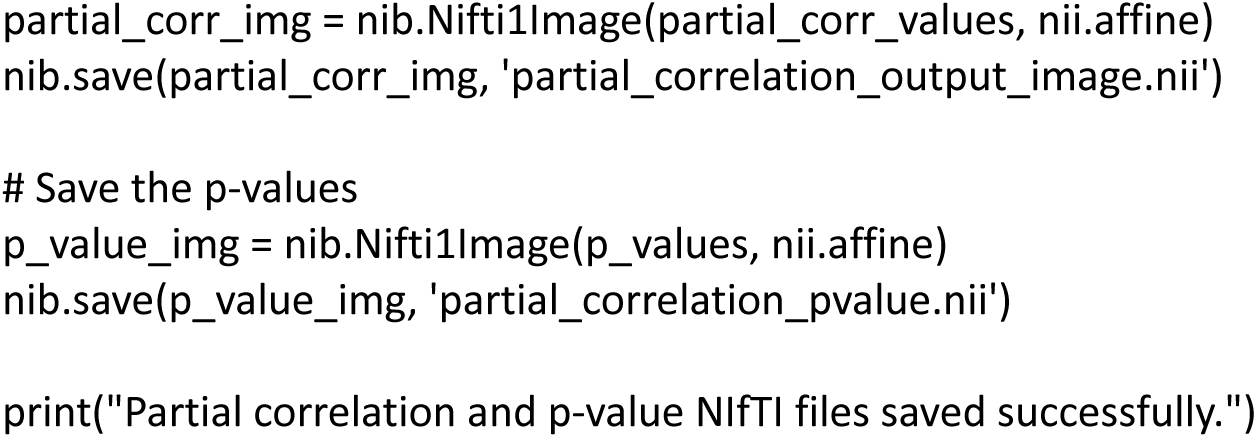

## Notes

### Competing Interest Statement

The authors have declared no competing interest.

### Author Declarations

The Office of Human Research Ethics at the University of North Carolina at Chapel Hill (UNC), the Veterans Affairs Puget Sound Health Care System Institutional Review Board, and the US Army Medical Research and Development Command Office of Human Research Oversight gave the ethical approvals for the human components of this work. The Veterans Affairs Puget Sound Health Care System's Institutional Animal Care and Use Committee (IACUC) gave ethical approval for animal components of this work.

